# What do the Welsh public understand about NHS dental services, what do they think they could look like, and what are their priorities? A qualitative study

**DOI:** 10.1101/2024.08.14.24311616

**Authors:** Natalie Joseph-Williams, Abubakar Sha’aban, Francesca Mazzaschi, Anthony Cope

**Affiliations:** Health and Care Research Wales Evidence Centre, Cardiff University, United Kingdom; Health and Care Research Wales Evidence Centre Public Partnership Group, Cardiff University, United Kingdom

## Abstract

NHS General Dental Services in Wales are undergoing reform. To ensure dental services meet the needs of those who use them, we explored what the public think these services could look like and what their priorities for are.

The aim of this study was to consult with the Welsh public to understand their views on NHS dental services to help inform dental reform plans in Wales. Specific objectives were to explore:

What do the public think Welsh NHS dental services could look like?

What do the public understand about dental services and the dental team?

What are their views on skill mix in dentistry?

What are their attitudes towards and needs for oral health self-management?

What are their priorities for dental care services in Wales?

We used qualitative methods (interviews and focus group style workshops) across two phases between November 2023 and May 2024 to explore the study objectives. Thematic analyses were performed on the data to identify key themes. Forty four participants with diverse backgrounds from all seven local health boards in Wales took part.

## 1. Background

> *“Having that public perspective on what is an acceptable service is absolutely vital when deciding on policy…any reform to services needs to reflect that public need and their preferences” Andrew Dickenson, Chief Dental Officer for Wales, 2024*

NHS General Dental Services in Wales are undergoing reform to a needs-based and preventive approach to care provision.[1] A recent report by The Health Foundation and Ipsos found that 60% of people would prioritise primary and community care (including dentistry) over care in hospitals.[2] This highlights the value that the increasing value the public place on these services. To ensure dental services meet the needs of those who use them, those who are involved in the redesign want to explore what the public think these services could look like and what their priorities for dental care are.

The Dental Contract Reform Programme was introduced by Welsh Government in 2017[1] as part of broader system reform in NHS General Dental Services in Wales. The dental contract reform programme acknowledges that Units of Dental Activity (UDA) as a sole measure of contract performance is not an ideal method of assessment, and no longer measures performance solely on these. Instead, the reform programme focuses on increasing *access* to dental services, improved delivery of *evidence-based prevention*, the implementation *of needs-led dental recall intervals*, and an *increase in the use of skill mix* (see Figure 1).

**Figure 1:**
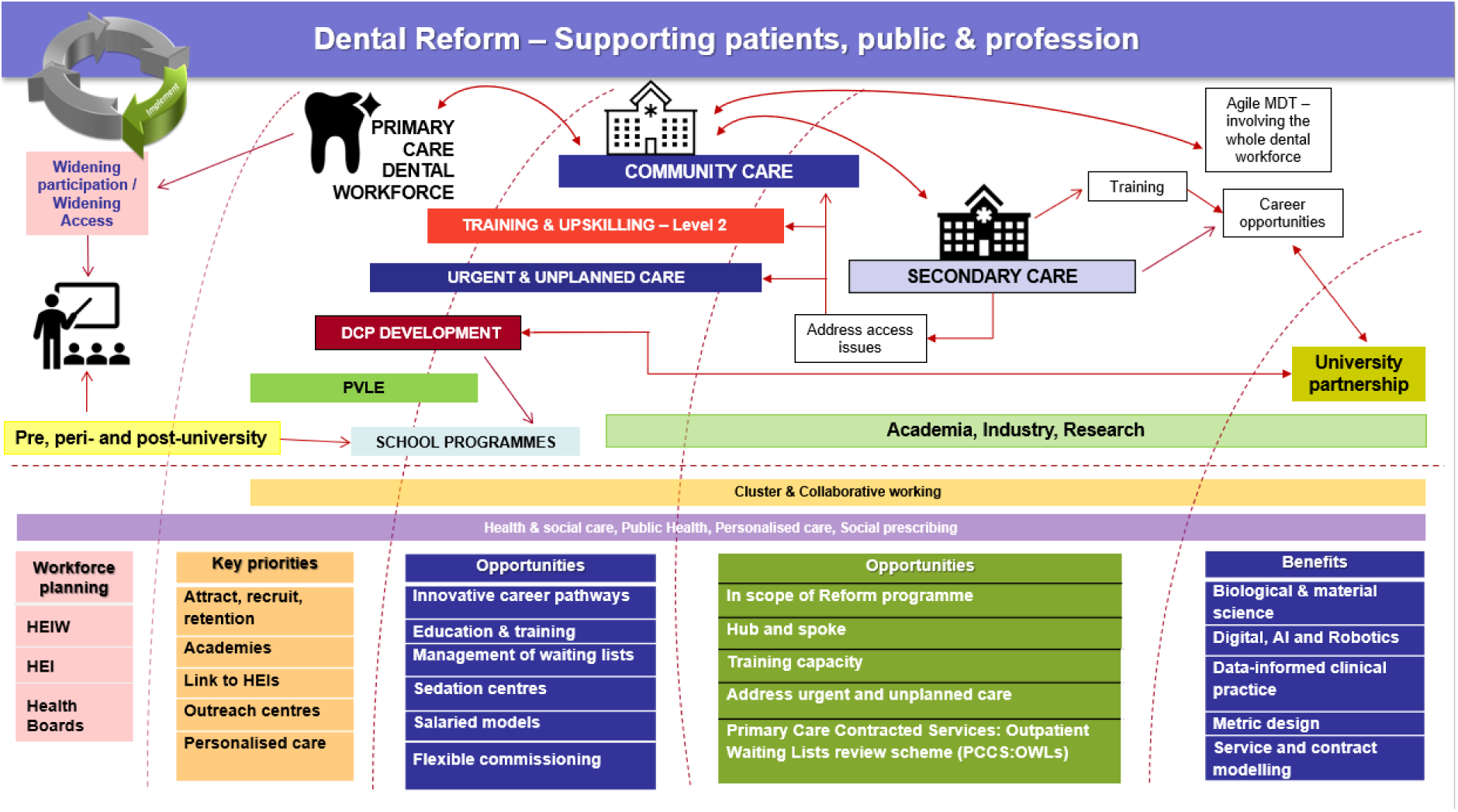
Overview of the dental reform programme in Wales

At the heart of the Dental Reform Programme are patients’ needs and preferences. Working with patients to discuss treatment options and co-produce care and recall plans means they will receive personalised care that aligns with the personal goals, which can lead to improved outcomes. However, moving beyond patient involvement in specific treatment and management decisions, it is also **necessary to explore patient’s preferences, needs, and priorities for what they think dental care in Wales could look like.**

Previous work has provided some insight into public impression and experiences of accessing and receiving dental services in Wales, including surveys[3, 4] and the collection of patient stories[5]. For example, stories collected by Public Health Wales[5] explored challenges of accessing NHS dental care, access during COVID-19 restrictions, past experiences, oral hygiene behaviour, and ways in which information on oral health is accessed. Whilst the results of the surveys and stories are important, they focus more on previous experiences and barriers to access, and less on what participants think and ideal dental service could look like.

This current study addresses this gap – exploring co-produced solutions to the reported challenges, which align with both the goals of the Dental Reform Programme and the needs of the people using these services. The results of this study will help to inform reform conversations in Wales, and support service planning that aligns with the key priorities of the Welsh population.

## 2. Aims and Objectives

- What do the public think Welsh NHS dental services could look like?
- What do the public understand about dental services and the dental team?
- What are their views on skill-mix in dentistry?
- What are their attitudes towards and needs for oral health self-management?
- What are their priorities for dental care services in Wales?

## 3. Methods

We used qualitative methods (interviews and focus group style workshops) across two phases to explore what the public think Welsh NHS dental services could look like, their understanding of the dental team, and what their priorities for dental care and services received are.

Phase 1 interviews focused on *what* people think NHS dental services should include, *how* it should be delivered, *who* should deliver the services, *what matters most* to people in terms of how and when they receive dental care, and their *understanding* of when and how to access *emergency dental care* (EDC). Interviews also explored preferences for being involved in the co-design of dental services.

Phase 2 workshop and interviews focused on peoples’ *understanding* of the *‘dental team’* and the services that they provide, *preferences* for which member of the team they see, *attitudes, barriers, facilitators* towards *skill mixing* and *multidisciplinary* approach, understanding of how and when to *access EDC, patient’s role* in their dental care and self-management, self-management *information needs,* and *what matters most* in terms of dental care and the services received.

Ethical approval for this study was granted by Cardiff University School of Medicine Research Ethics Committee (SMREC 23.71). Recruitment processes and documents were developed with our Public Partner (AC) to ensure they were feasible, understandable, and relevant.

### 3.1 Participant recruitment

#### 3.1.1 Phase 1 interviews

Included participants were people eligible to access NHS General Dental Services (GDS) in Wales and were 18 years of age or over at the time of the interview. Participants were recruited through social media using ‘X’ (formally known as Twitter) accounts of using Health and Care Research Wales, Health and Care Research Wales Evidence Centre and The Wales Centre for Primary and Emergency Care Research (PRIME Centre Wales) accounts.

The opportunity was also shared through other relevant email shared lists (e.g. PRIME Centre Wales). Advertisements included a poster summarising the study (Appendix A) and a link to the study webpage, which included a detailed participant information sheet (Appendix B).

Interested participants were asked to complete an online screening and demographic survey (Appendix C) to register their interest in taking part. Demographic variables collected included: gender, age group, health board area, current NHS dental registration status, disability status, highest educational level, employment status, personal income range (previous year), and ethnicity. We also asked participants three questions exploring components of digital literacy[6]. We used data from the screening and demographic survey to purposively select a diverse and representative sample. If selected, participants were notified via email and asked to complete a consent form (Appendix D); a convenient date and time were agreed upon for the interview with the researcher (AS/FM).

#### 3.1.2 Phase 2 workshop and interviews

Included participants were people eligible to access NHS General Dental Services (GDS) in Wales and were 18 years of age or over at the time of the workshop/interview. Using a word of mouth approach, the opportunity was advertised using a poster (Appendix E) through various internal and external networks (including mailing lists and the Health and Care Research Wales Evidence Centre Public Partnership Group). Participants could view the participant information sheet via the website link provided (Appendix F for workshop, Appendix B for phase 2 interviews). As mentioned above, interested participants were invited to complete an online screening and demographic survey (Appendix C). A purposive sample was invited to participate and complete a brief 5-minute consent discussion via phone or Microsoft Teams and complete a consent form (Appendix G for workshop, Appendix D for phase 2 interviews). Participants were sent log in details for the online workshop. For interviews, a convenient date and time were agreed for the interview with the researcher (AS).

### 3.2 Data collection

#### 3.2.1 Phase 1 interviews

Semi-structured interviews were conducted online via Microsoft Teams between November 2023 and January 2024. Interviews were conducted by AS or FM and audio/video recorded. To help aid interview discussions, participants were sent a brief information sheet prior to the interview, outlining definitions of non-urgent, urgent, and emergency dental services (Appendix H). Researchers (AS/FM) used an interview schedule (Appendix I) to guide the discussion and ensure key topics were explored. This was developed with stakeholders, including our Public Partner (AC), to ensure the questions were accurate, accessible, and covered areas that mattered to the public. A summary of key topics covered in Phase 1 is included in Box 1.

###### Box 1: Summary of key topics explored during Phase 1 Interviews

***What*** should general dental care in Wales include (how to access, what they expect when accessing)?

***How*** should general dental care be delivered in Wales?

***Who*** should be involved in delivery of general dental care? (Including current understanding of who is in the dental team and attitudes towards, preferences for, and barriers/facilitators to seeing different members of the dental team)

What do the public understand about emergency dental care?

***What matters most to dental patients*** in terms of how and when they receive dental care?

#### 3.2.2 Phase 2 workshop and interviews

A workshop was conducted in April 2025 via Zoom (led by NJW). Phase 2 interviews were conducted in May 2025 via Teams. The workshop and interviews were audio/video recorded for transcription. The workshop guide (Appendix J) provides details of how the 1.5 hour workshop was run. The slides used in the workshop are included in Appendix K. Key topics discussed during the workshop and Phase 2 interviews are include in Box 2.

###### Box 2: Summary of key topics explored during Phase 2 workshop and interviews

**Topic 1 – What do you understand about the ‘dental team’ & the services they provide?**

- Who is part of the *dental team*?
- *Understanding* of the different team members’ *roles*
- What would help to *improve understanding*?
- *Preferences* for dental team members
- Attitudes, barriers, and facilitators towards *skill-mixing* and *multidisciplinary* approach to GDS

**Topic 2 – Dental care services**

- *how* and *when* to access *emergency dental care*
- what *role patients should play* in their own dental care
- self-management experiences and *information/support needs*

**Topic 3 – What matters most to you?**

- *What matters most to you* in your dental care and the services you receive?
- What would be your *number one thing* that must be achieved in an *ideal and acceptable NHS dental service?*

### 3.3 Data Analysis

Phase 1 interviews and Phase 2 workshop and interviews were transcribed verbatim for analysis using a Cardiff University authorised third-party transcription service. Transcripts were imported into NVivo 12 pro Qualitative Analysis Software[7] for analysis. Thematic analysis[8] was applied separately to Phase 1 and Phase 2 data[9] by AS and FM; this is a systematic method used to identify patterns and themes within data. It involves a series of structured steps including familiarisation with the data, coding, developing a thematic framework, charting the data into the framework (including verbatim quotes), and interpreting the key themes.

The initial thematic coding framework (emerging themes from preliminary exploration of the data) were applied to a set of transcripts (n=6) by FM for the phase 1 data and AS for the phase 2 data and cross-checked by AS and FM respectively. The emerging frameworks (Phase 1 and Phase 2) were discussed with NJW and refined (similar codes merged and additional codes added to the framework) before applying the frameworks to the remainder of the transcripts.

Phase 1 and Phase 2 data were analysed separately in the first instance. However, where there were commonalities in themes emerging from Phase 1 and Phase 2 data, these were synthesised and are presented together in the key findings section (see section 4.2); for example, ‘what matters most’ was explored and identified as a key theme for both Phase 1 and Phase 2 data.

## 4. Results

### 4.1 Participants

We recruited a total of 44 participants across Phase 1 and Phase 2: 29 participants for Phase 1 interviews and 15 participants for Phase 2 (9 for the workshop and 6 for interviews).

Overall, 39% (n=17) of participants identified as female, 57% identified as male (n=25), and 4% identified as non-binary (n=2). Participants included in Phase 1 and Phase 2 reported a range of disability status, educational attainment, employment status, and personal income range (see Table 1).

**Table 1:** Demographic characteristics of Phase 1 and Phase 2 participants (n=44)

| Research ID | Study Phase | Gender<br>M = Male<br>F - Female | Age Range<br>(years) | Current NHS dental registration status | Local health board area | Disability Status | Highest educational level attained | Employment status | Personal income last year (£) | Ethnicity |
| --- | --- | --- | --- | --- | --- | --- | --- | --- | --- | --- |
| **Phase 1 Interviews (n = 29)** |  |  |  |  |  |  |  |  |  |  |
| P1 | Phase 1 interview | M | 25 – 34 | Not sure | Cardiff and Vale University Health Board | Chronic illness | University level qualification | Self-employed | 1 to 9, 999 | Asian or Asian British |
| P3 | Phase 1 interview | F | 65 – 74 | Currently registered with an NHS dentist | Hywel Dda University Health Board | None | University level qualification | Retired | 10, 000 to 24, 999 | White |
| P4 | Phase 1 interview | F | 45 – 54 | Currently registered with an NHS dentist | Aneurin Bevan University Health Board | Neurodivergent (e.g., autism, ADHD) | University level qualification | Employed part-time | 25, 000 to 49, 999 | White |
| P5 | Phase 1 interview | F | 25 – 34 | Currently registered with an NHS dentist | Aneurin Bevan University Health Board | None | University level qualification | Student (full-time) | 10, 000 to 24, 999 | White |
| P6 | Phase 1 interview | F | 45 – 54 | Not currently registered with an NHS dentist | Cardiff and Vale University Health Board | Chronic illness | University level qualification | Employed full-time | 75, 000 to 99, 999 | White |
| P7 | Phase 1 interview | M | 25 – 34 | Currently registered with an NHS dentist | Cardiff and Vale University Health Board | None | University level qualification | Employed full-time | 25, 000 to 49, 999 | White |
| P8 | Phase 1 interview | F | 65 – 74 | Currently registered with an NHS dentist | Betsi Cadwaladr University Health Board | None | University level qualification | Self-employed | 10, 000 to 24, 999 | White |
| P9 | Phase 1 interview | F | 55 – 64 | Currently registered with an NHS dentist | Betsi Cadwaladr University Health Board | None | University level qualification | Retired | Prefer not to say | White |
| P12 | Phase 1 interview | M | 25 – 34 | Currently registered with an NHS dentist | Cwm Taf Morgannwg University Health Board | None | University level qualification | Employed full-time | 10, 000 to 24, 999 | Black, African, Caribbean or Black British |
| P13 | Phase 1 interview | M | 35 – 44 | Not currently registered with an NHS dentist | Cardiff and Vale University Health Board | Mobility impairment | University level qualification | Self-employed | 10, 000 to 24, 999 | Mixed or Multiple ethnic groups |
| P18 | Phase 1 interview | M | 25 – 34 | Currently registered with an NHS dentist | Swansea Bay University Health Board | None | University level qualification | Employed part-time | 10, 000 to 24, 999 | White |
| P22 | Phase 1 interview | M | 75 – 84 | Currently registered with an NHS dentist | Betsi Cadwaladr University Health Board | Hearing impairment | School or college leaver qualification | Retired | 10, 000 to 24, 999 | White |
| P23 | Phase 1 interview | M | 25 – 34 | Currently registered with an NHS dentist | Cwm Taf Morgannwg University Health Board | Chronic illness | University level qualification | Employed part-time | 25, 000 to 49, 999 | Black, African, Caribbean or Black British |
| P24 | Phase 1 interview | M | 55 – 64 | Not sure | Betsi Cadwaladr University Health Board | Stroke survivor - some anxiety, mental health issues and concentration difficulties | University level qualification | Retired early due to stroke, unable to work | 25, 000 to 49, 999 | White |
| P27 | Phase 1 interview | M | 25 – 34 | Currently registered with an NHS dentist | Swansea Bay University Health Board | None | University level qualification | Employed full-time | 50, 000 to 74, 999 | Black, African, Caribbean or Black British |
| P28 | Phase 1 interview | M | 25 – 34 | Currently registered with an NHS dentist | Swansea Bay University Health Board | None | School or college leaver qualification | Employed part-time | 25, 000 to 49, 999 | Black, African, Caribbean or Black British |
| P30 | Phase 1 interview | M | 18 – 24 | Currently registered with an NHS dentist | Swansea Bay University Health Board | None | University level qualification | Student (full-time) | 25, 000 to 49, 999 | White |
| P33 | Phase 1 interview | M | 18 – 24 | Currently registered with an NHS dentist | Aneurin Bevan University Health Board | None | School or college leaver qualification | Employed part-time | 10, 000 to 24, 999 | Black, African, Caribbean or Black British |
| P35 | Phase 1 interview | M | 25 – 34 | Currently registered with an NHS dentist | Betsi Cadwaladr University Health Board | None | School or college leaver qualification | Employed full-time | 75, 000 to 99, 999 | White |
| P36 | Phase 1 interview | M | 18 – 24 | Currently registered with an NHS dentist | Swansea Bay University Health Board | None | School or college leaver qualification | Employed part-time | 25, 000 to 49, 999 | Black, African, Caribbean or Black British |
| P37 | Phase 1 interview | F | 25 – 34 | Currently registered with an NHS dentist | Not Applicable | None | University level qualification | Employed part-time | 25, 000 to 49, 999 | Black, African, Caribbean or Black British |
| P38 | Phase 1 interview | F | 18 – 24 | Currently registered with an NHS dentist | Powys Teaching Health Board | Visual impairment | School or college leaver qualification | Self-employed | 25, 000 to 49, 999 | Black, African, Caribbean or Black British |
| P43 | Phase 1 interview | F | 45 – 54 | Currently registered with an NHS dentist | Not sure | Mobility impairment | No formal qualifications | Unable to work due to illness or disability | 10, 000 to 24, 999 | White |
| P44 | Phase 1 interview | M | 25 – 34 | Not currently registered with an NHS dentist | Powys Teaching Health Board | Mental health condition | University level qualification | Employed part-time | 50, 000 to 74, 999 | Black, African, Caribbean or Black British |
| P45 | Phase 1 interview | F | 35 – 44 | Not sure | Cardiff and Vale University Health Board | Neurodivergent (e.g., autism, ADHD) | School or college leaver qualification | Unable to work due to illness or disability | 1 to 9, 999 | White |
| P49 | Phase 1 interview | F | 25 – 34 | Currently registered with an NHS dentist | Cardiff and Vale University Health Board | None | School or college leaver qualification | Employed part-time | 50, 000 to 74, 999 | Black, African, Caribbean or Black British |
| P52 | Phase 1 interview | M | 25 – 34 | Currently registered with an NHS dentist | Cardiff and Vale University Health Board | None | University level qualification | Employed full-time | 50, 000 to 74, 999 | Mixed or Multiple ethnic groups |
| P53 | Phase 1 interview | F | 25 – 34 | Currently registered with an NHS dentist | Cardiff and Vale University Health Board | None | University level qualification | Employed part-time | 50, 000 to 74, 999 | Black, African, Caribbean or Black British |
| P55 | Phase 1 interview | F | 45 – 54 | Currently registered with an NHS dentist | Aneurin Bevan University Health Board | None | School or college leaver qualification | Retired | 100, 000 or more | White |
| **Phase 2 Workshops and Interviews (n=15)** |  |  |  |  |  |  |  |  |  |  |
| P2 | Phase 2 Workshops | M | 55 – 64 | Currently registered with an NHS dentist | Aneurin Bevan University Health Board | None | University level qualification | Retired | £10, 000 to £24, 999 | White |
| P21 | Phase 2 Workshops | M | 65 – 74 | Currently registered with an NHS dentist | Cardiff and Vale University Health Board | None | University level qualification | Prefer not to say | £10, 000 to £24, 999 | White |
| P25 | Phase 2 Workshops | M | 65 – 74 | Currently registered with an NHS dentist | Cardiff and Vale University Health Board | None | University level qualification | Retired | £25, 000 to £49, 999 | White |
| P88 | Phase 2 Workshops | M | 25 – 34 | Currently registered with an NHS dentist | Aneurin Bevan University Health Board | None | University level qualification | Employed full-time | £25, 000 to £49, 999 | Black, African, Caribbean or Black British (African background) |
| P122 | Phase 2 Workshops | M | 25 – 34 | Currently registered with an NHS dentist | Cardiff and Vale University Health Board | None | University level qualification | Employed full-time | £50, 000 to £74, 999 | White |
| P134 | Phase 2 Workshops | non-binary | 25 – 34 | Currently registered with an NHS dentist | Swansea Bay University Health Board | None | University level qualification | Self-employed | £10, 000 to £24, 999 | Mixed or Multiple ethnic groups (White and Black Caribbean) |
| P104 | Phase 2 Workshops | non-binary | 25 – 34 | Currently registered with an NHS dentist | Swansea Bay University Health Board | None | University level qualification | Self-employed | £25, 000 to £49, 999 | Prefer not to say |
| P174 | Phase 2 Workshops | M | 35 – 44 | Not currently registered with an NHS dentist | Swansea Bay University Health Board | None | University level qualification | Employed full-time | £50, 000 to £74, 999 | White |
| P173 | Phase 2 Workshops | F | 65 – 74 | Not currently registered with an NHS dentist | Swansea Bay University Health Board | None | School or college leaver qualification | Employed part-time | £10, 000 to £24, 999 | White |
| P71 | Phase 2 interview | M | 45 – 54 | Currently registered with an NHS dentist | Hywel Dda University Health Board | None | University level qualification | Employed full-time | Prefer not to say | White |
| P72 | Phase 2 interview | M | 75 – 84 | Not currently registered with an NHS dentist | Aneurin Bevan University Health Board | Type 1 Diabetes | School or college leaver qualification | Retired | £10, 000 to £24, 999 | White |
| P73 | Phase 2 interview | F | 45 – 54 | Currently registered with an NHS dentist | Powys Teaching Health Board | Hearing impairment | University level qualification | Employed full-time | £25, 000 to £49, 999 | White |
| P74 | Phase 2 interview | F | 55 – 64 | Currently registered with an NHS dentist | Cardiff and Vale University Health Board | None | University level qualification | Employed part-time | £1 to £9, 999 | White |
| P75 | Phase 2 interview | M | 45 – 54 | Currently registered with an NHS dentist | Cardiff and Vale University Health Board | None | School or college leaver qualification | Prefer not to say | Prefer not to say | Mixed ethnicity |
| P76 | Phase 2 interview | F | 35 – 44 | Not currently registered with an NHS dentist | Swansea Bay University Health Board | None | University level qualification | Employed full-time | £25, 000 to £49, 999 | White |

#### 4.1.1 Local Health Board Area

All seven health boards in Wales were represented by participants across Phase 1 and Phase 2 combined (see Figure 2); overall, 30% (n=13) of participants lived in the Cardiff & Vale University Health Board area. All seven health boards were represented in Phase 1. Five of the seven health boards were represented in Phase 2; Betsi Cadwaladr and Cwm Taf Morgannwg were not represented.

**Figure 2:**
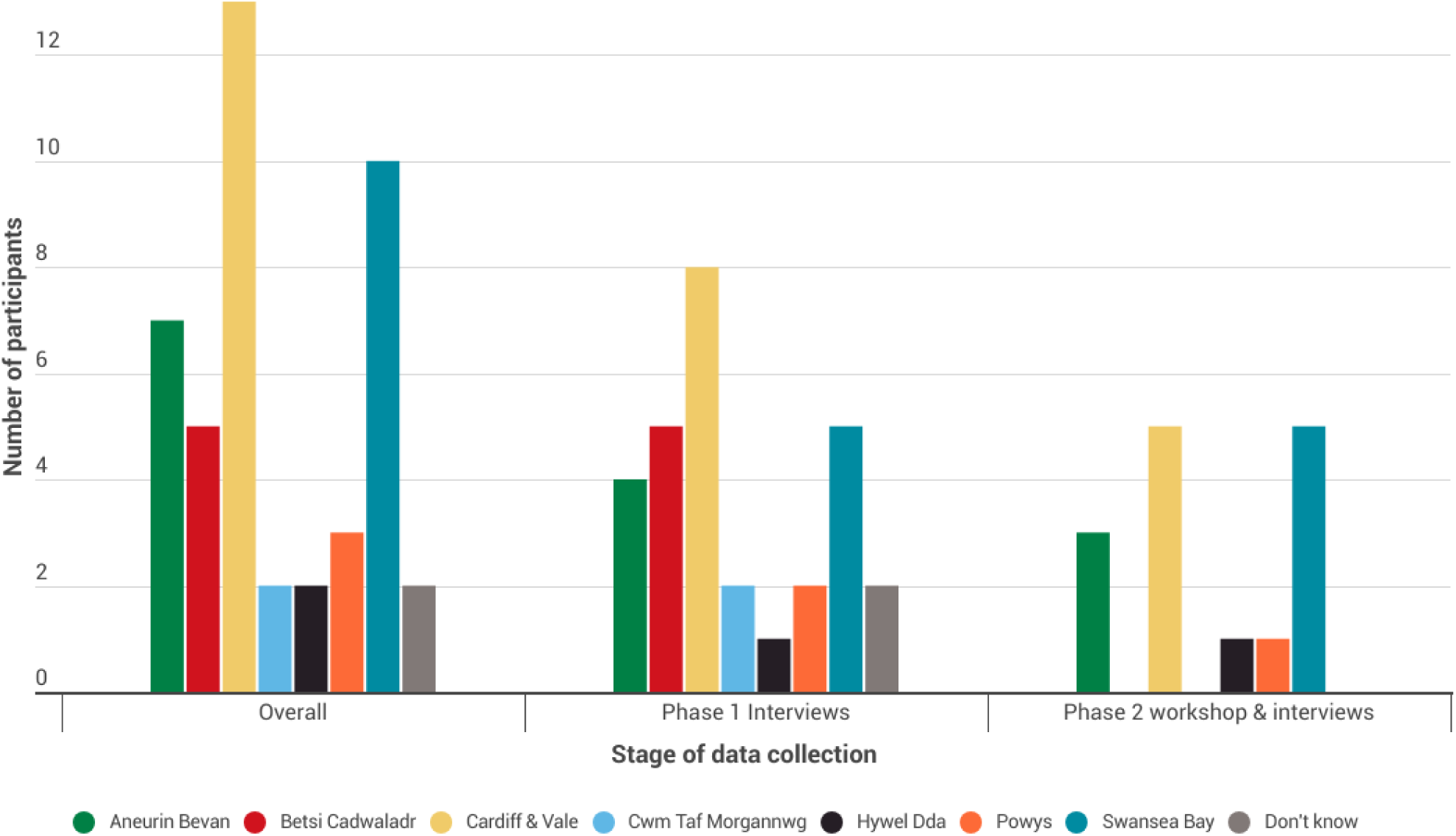
Local health Board area of participants: Overall, Phase 1 and Phase 2

#### 4.1.2 NHS dental registration

Current NHS dental registration of participants across Phase 1 and 2 combined included:

- 34 (77%) *currently registered* with an NHS dentist
- 7 (16%) *not currently registered* with an NHS dentist
- 3 (7%) *unsure* if they are registered with an NHS dentist

#### 4.1.3 Ethnicity

A range of ethnicities were reported by the 44 participants included in Phase 1 and Phase 2 (see Figure 3). Overall, participants report their ethnicity as: White (n=26); Black, African, Caribbean, or Black British (n=12); Mixed or multiple ethnic groups (n=4); Asian or Asian British (n=1); or prefer not to say (n=1).

**Figure 3:**
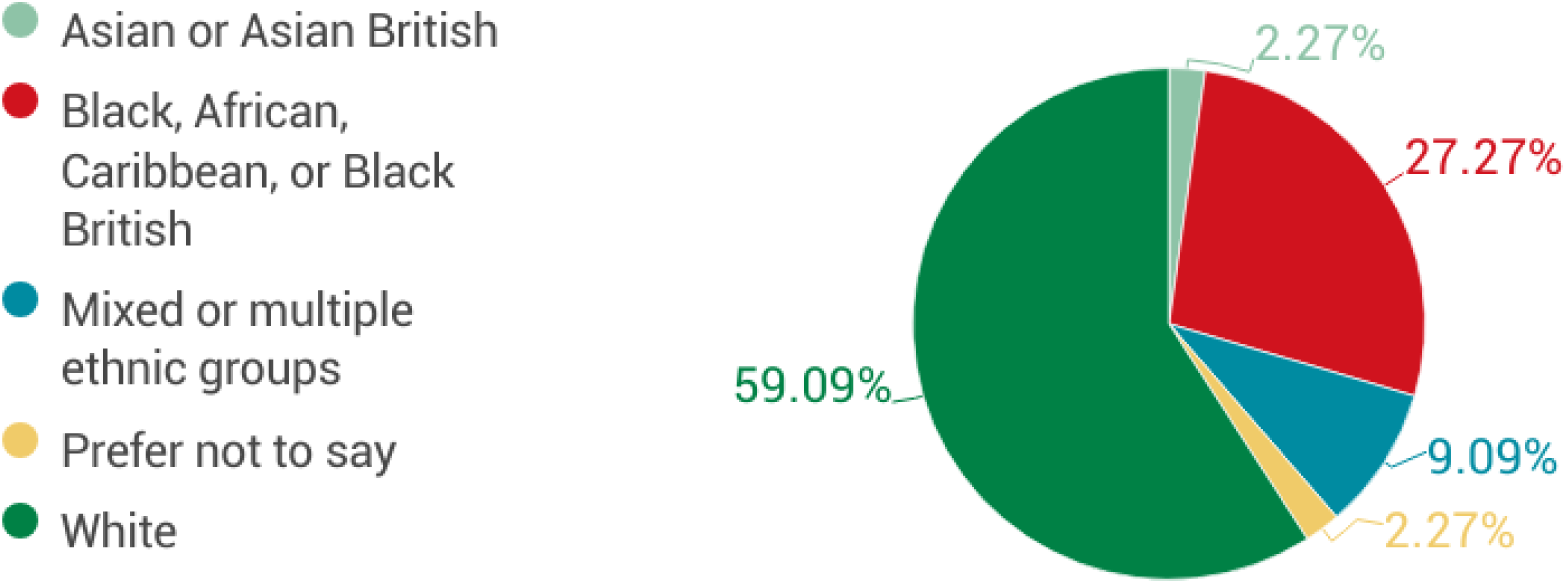
Percentage of participants in each ethnic group (Phase 1 and Phase 2)

#### 4.1.4 Age

Participants included in Phase 1 and Phase 2 (n=44) ranged in age category from 18-24 to 75-84; the most frequent age category was 25-34 (see Figure 4).

**Figure 4:**
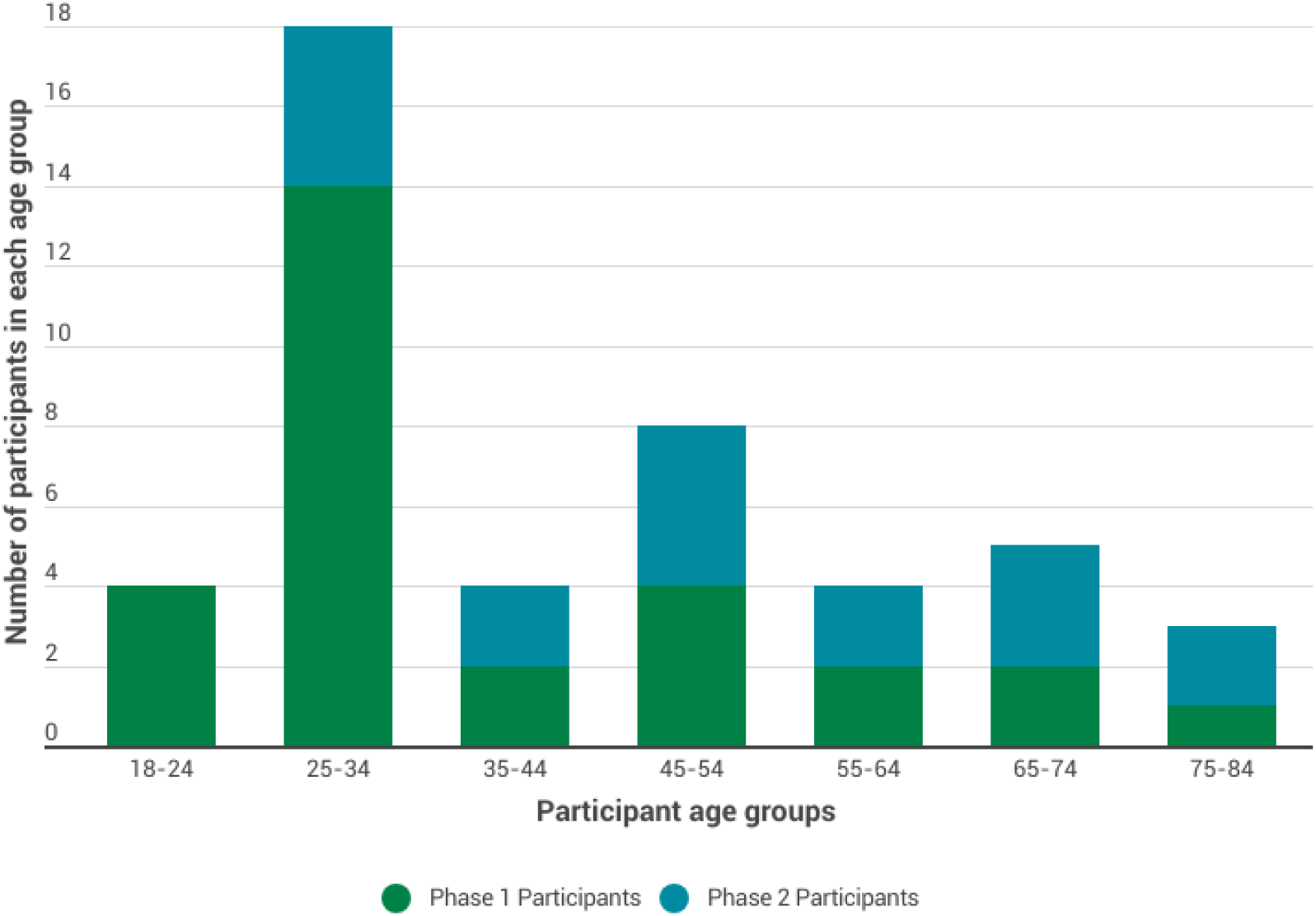
Age categories of Phase 1 and Phase 2 participants

#### 4.1.5 Digital Literacy

Participants reported a range of self-reported ‘digital literacy’, covering the use of applications, setting up video calls, and solving basic technical issues (see Figure 5).

**Figure 5:**
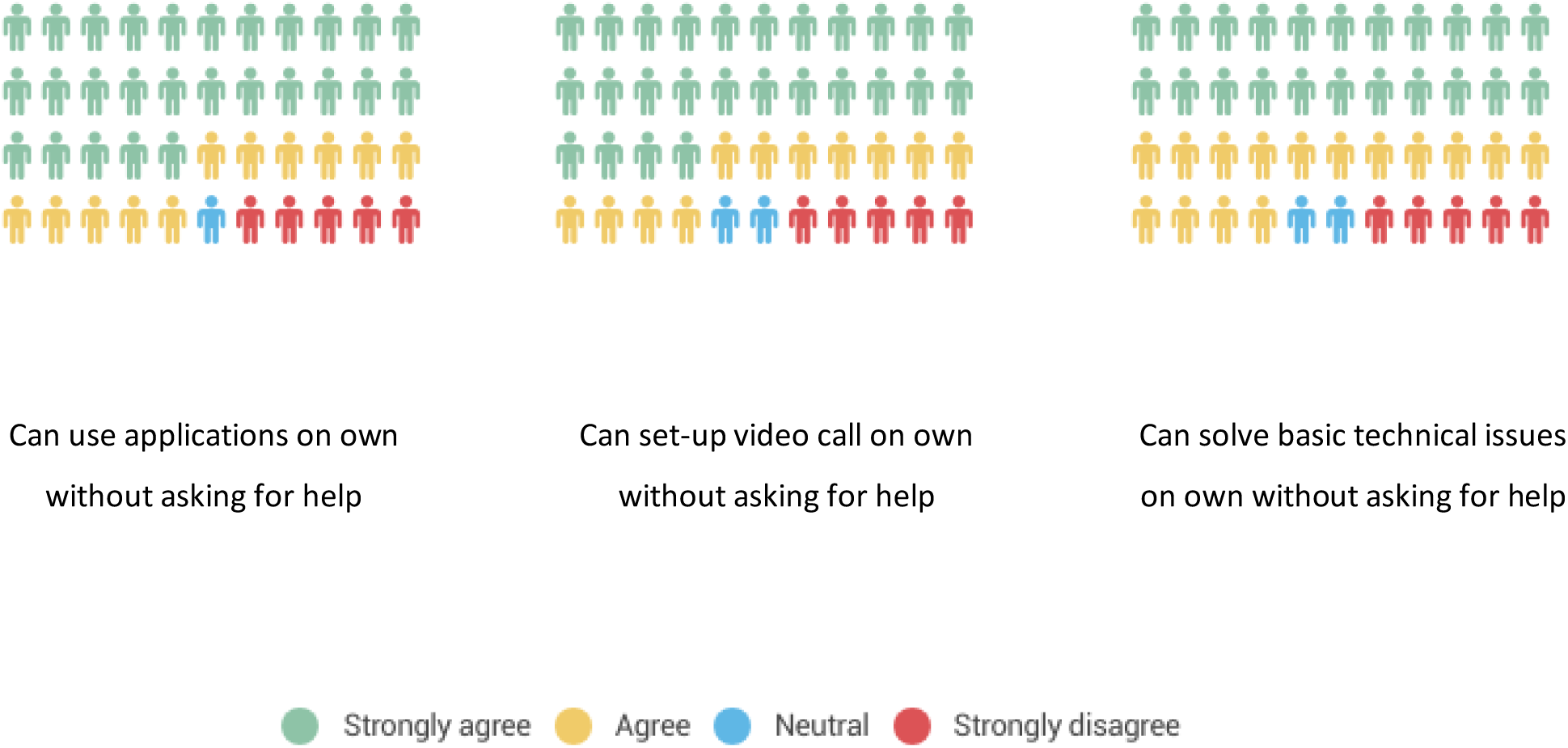
Self-reported level of digital literacy (Phase 1 and 2 participants combined)

##### Use of applications

Overall, 38 participants (86%) agreed or strongly agreed that they could use applications/programs (like Zoom) on their mobile phone, computer, or another electronic device (for example a tablet) on their own without asking for help from others. Five participants strongly disagreed, and one participant was neutral.

##### Setting up video calls

Overall, 37 participants (84%) agreed or strongly agreed that they could set up video calls on their mobile phone, computer, or another electronic device on their own without asking for help from others. Five participants strongly disagreed and two were neutral.

##### Solving basic technical issues

Overall, 37 participants (84%) agreed or strongly agreed that they could solve or figure out how to solve basic technical issues on their own without asking for help from others. Five participants strongly disagreed and two were neutral.

### 4.2 Key findings

Overall, **seven key themes emerged across both data sets** (Phase 1 interviews and Phase 2 Workshop and Interviews). Key themes are summarised in Table 2. Key themes and sub- themes are presented in Additional Table A (separate file) with exemplar quotes^1^, and are presented in turn below.

**Table 2.** Summary of key themes.

| **Theme** | **Report Section** | **Page Number** |
| --- | --- | --- |
| **Understanding of the ‘dental team’ and their roles** | 4.2.1 | 24 |
| **Skill-mix and a whole-team approach to dental service delivery** | 4.2.2 | 26 |
| **Urgent and emergency dental services** | 4.2.3 | 30 |
| **Essential general dental services and mode of delivery** | 4.2.4 | 32 |
| **Digital technology and ‘teledentistry’** | 4.2.5 | 35 |
| **Self-management of oral health** | 4.2.6 | 38 |
| **What matters most to the public – key priorities for dental services in Wales** | 4.2.7 | 46 |

#### 4.2.1 Understanding of the ‘dental team’ and their roles

This theme emerged from Phase 1 and Phase 2 data. Participants reflected on their understanding of the dental team and their roles and how this compares to understanding of other healthcare teams. They also made suggestions for how dental team understanding could be improved.

Prior to data collection, we sought clarification on the primary care dental team roles as a guide to assess participants’ awareness. Dental team roles include dentist, dental nurse, hygienist, dental therapist, dental technician, and orthodontist (see Appendix L for role descriptions).

##### 4.2.1.1 Awareness of the ‘dental team’

Overall, the **most frequently cited** primary care dental team **roles were the dentist, dental nurse, and hygienist:**

> *“I could only identify three roles. One being the receptionist, the other being the dentists themselves, and then the third one being the oral hygienist or something like that. I think those are the only three roles that I could actually identify in there.”* P08 Phase 1

> *“The dentist, the hygienist, the dental nurse, the receptionist, the, and then whatever specialty you get referred to as well.”* P45 Phase 1

During Phase 1 interviews, eleven participants reported **limited understanding of the dental team beyond the dentist:**

> *“I heard about the dentist, it’s all about the dentists to me.”* P33 Phase 1

> *“So, I feel apart from a dentist, the dental specialist. I don’t know. It is your dentist, and the dental specialist should be able to look after my dental care.”* P35 Phase 1

Only a **limited number of participants referred to the other roles** that are part of the primary care dental team (see Appendix L): ‘*dental technicians’* (one participant) and ‘*orthodontists’* (two participants). No participants reported being aware of ‘*dental therapists’*. Some participants referred to ‘dental surgeons’ and ‘specialists’ but recognised that they would provide care elsewhere and likely require a referral.

Whilst not specifically defined as a primary care dental role (see Appendix L), **many participants referred to the role of ‘dental assistant’.** This term was typically used to identify anyone (regardless of their qualified role) who provided a supporting role to the dentist:

> *“So thinking about the dental team, I would, I would usually refer to the dentist, who is really a core part of the dental team. And also there is the dental assistants that help the dentist when he is about to perform a procedure.”* P4 Phase 2

> *“Okay. As far as I’m aware, there’s the, the big man at the top [dentist’s name] who owns the practice. He’s got a team of dentists who work underneath him. They’ve got a couple of hygienists, and they’ve got assistants.”* P22 Phase 1

Most participants seemed **aware that there were different roles performed by the other team members** and described tasks that would be done by a hygienist (e.g. scale and polish), a therapist (e.g. x-rays or teeth impressions) or technician (e.g. making bridges or dentures), **but they were uncertain who performed them and what they were called;** often reverting to the term ‘dental assistant’.

> *“Okay, I think the dental team consists of a couple of, number of people ranging from the dentist and, and the hygienist and we do not know how to name them, but I know that they have different roles. Yeah, they do not do the same thing.”* P37 Phase 1

> *P3: My understanding is the dental assistant can either be the dental technician or the dental nurse. I think it depends on the practice.”* P34 Phase 2

One participant **did not feel that they had different titles and provide different services**, instead, seeing them all as ‘dentists’ and as **part of ‘one family’:**

> *“They’re still dentists, they’re still part of the dental services. So although you’ve listed them out separately, for me, and for most laypeople, they see them as one family…they don’t see them as different…they might be different individuals, but they’re not different service care providers.”* P75 Phase 2

Whilst not formally included as a primary care dental team role (see Appendix L), **many participants perceived administrative staff as a core part of the ‘dental team’.** They coordinate appointments, help patients to navigate their care, and are a key point for communication with patients and between professionals.

> *“So this and also the receptionist…I would regard them as, you know, a core member of the team. Because they are the ones responsible for receiving patients and then communication with the dentist.”* P134 Phase 2

> *“I’d include the receptionist, as part of the team, because there’s a lot they can reassure people on and spot when someone comes in. Obviously, the main provider is the dentist himself or herself, but [they] are an important part of that as well.”* P24 Phase 1

One participant felt that **patients were also a core part of the ‘dental team’:**

> *“I mean, those who are involved, like the patients, the community support groups. So, to me, when we talk about teams, it shall be not just the service providers, but the service users as well.”* P75 Phase 2

##### 4.2.1.2 Less understanding of the ‘dental team’ compared to other health care teams

**Some participants reflected that they had** less understanding of dental teams compared to other health care teams **that they interact with:**

> “I would say I have a much greater understanding of other teams than a dental team. I didn’t realise there was so much available to a dental team because I’ve never been offered it. And I’ve never experienced it. And I’ve never noticed these other positions exist.” P73 Phase 2

> “I definitely have less understanding of what a dental team comprises of and their roles, compared to like you say, a GP practice. I’d have far more understanding if I was to go to a GP practice and they said “you’re best off speaking to the physiotherapist who comes here once a week rather than the GP”. Whereas, a kind of similar example for the dentist I wouldn’t I know why or who I’d be seeing and why I wouldn’t be seeing the dentist.” P174 Phase 2

One participant reported that they were **unclear how dental team members work together:**

> “If I visited my GP surgery, and I saw, for example, a nurse practitioner…or a phlebotomist…then I would absolutely know that if they had concerns or needed to check something that they were going to be going off to the doctor, having their meetings, deciding what the best way forward is. But because I guess they’re working together, they’re in the same building, they might even just call their colleague in. Whereas in the dental world, I suppose I don’t have so much of that picture.” P21 Phase 2

#### 4.2.2 Skill-mix and a whole-team approach to dental service delivery

This theme emerged from Phase 1 and Phase 2 data. Participants reflected on their preferences for consulting with the different team members and whether they would be happy to receive care from members of the dental team other than the dentist.

##### 4.2.2.1 Dental team member preference

Overall, most participants reported that they would be happy to see other members of the dental team for issues they would typically see the dentist for. The predominant view was that **it was less important ‘who’ they saw – what mattered most was seeing the right person for the issue.** If the other team members are qualified and perceived as competent to provide the care, patients are positive towards skill-mixing in their dental care:

> *“I’d be happy enough to meet with whoever could give me the information that I wanted at the time… all I’m interested in is getting the right results.”* P8 Phase 1

> *“I don’t mind who I see as long as they’re qualified for what needs doing, you know?”* P3 Phase 1

> *“No, whoever’s qualified to do so. I don’t mind at all…whoever is competent at the end of the day.”* P4 Phase 1

A few participants **compared this approach to what they are already used to and comfortable with in other areas of health care:**

> *“I wouldn’t have a problem at all. I think we’ve done it with medicine, haven’t we? We’ve got nurse practitioners and other specialist areas, so to free up time with the doctors. I’ve got no issue at all doing that, seeing someone different in the dentist surgery if I needed to.”* P6 Phase 1

> *“I’d be completely relaxed about it being more akin to the sort of GP model where you’re going to see a nurse or a phlebotomist or a therapist or a GP. I’m sure there’s plenty of dental procedures that a dental nurse would be perfectly well qualified to perform, probably better than a dentist.”* P24 Phase 1

Some participants noted that **improved awareness and education is required**, so that people are aware that other dental team members are available, and they are **educated about the different roles and expertise:**

> *“…sometimes at the doctor’s, it’s very appropriate to see a nurse, a physiotherapist. You don’t always have to see the GP…if we were properly educated as to who can do what, and then we think ‘oh, well, that’s okay, I’m seeing the dental nurse. I’m not actually seeing the dentist today, but dental nurses know about and do this.”* P74 Phase 2

> *“…it’s kind of all tied together really, you know, making people really aware of what these different roles do and their expertise…they might have been through the same kind of training, and letting people know that.”* P174 Phase 2

However, there were a small number of participants who stated that they would only wish to see the dentist for anything other than cleaning procedures:

> *“But I’m sure I’m not alone in thinking when I go to the dentist, I’m gonna see the dentist… I know a lot of people, and I think it’s partly generational, that I always expect to see the GP about anything. and feel short-changed if it just happens to be a nurse.”* P24 Phase 1

> *“I mean, ultimately, my preference would be the dentist himself or herself. Because they’ve obviously got that all-encompassing knowledge.”* P73 Phase 1

##### 4.2.2.2 Potential benefits of skill-mixing and a whole-team approach

Participants discussed **the potential benefits of skill-mixing** and whole-team approaches to their dental care, where other team members take on more of the work that patient’s typically consult dentists for.

One potential benefit was fostering **patient choice** and **dental care that aligns with** patients’ preferences:

> *“Again, I say if they focus on the individual’s need and individual’s care then it shouldn’t be a problem. If those people are available, then you’ve got the choice. And I think that’s important. You give the individual a choice of what they want done and who they want to see.”* P72 Phase 2

Other participants noted that when **professionals work together**, this results in better outcomes for patients and provides a **more holistic service:**

> *“I think from the patient perspective, if a group of professionals are deciding together what is best for your care and keeping you informed, then that’s probably better for you…so you feel that you’re being well supported and you’re more likely to stick to what you need to stick to if all the right people are talking to you and talking to each other.”* P74 Phase 1

> *“You get potentially more of a holistic care because you get people who’ve got slightly different skills providing your care.”* P76 Phase 2

Some participants felt that a whole-team approach to dental care would also result in benefits for the dental team; this included **upskilling other members of the workforce** and **increasing moral and motivation to join the workforce:**

> *“I think, I think there’s a lot of benefits…for the team themselves, they may well have the opportunity to get up-skilled and carry out procedures that they might not have done before. I think there’s that kind of opportunity for sharing roles and responsibilities.”* P71 Phase 2

> *“…dentists are a bit on their knees as a profession, aren’t they? So, if they enjoy being in a multidisciplinary team and they’re all talking to each other and they’re getting a professional buzz out of it, then that might encourage more to come into the profession, which is obviously what we need.”* P74 Phase 2

Practically, participants felt that this approach could **free up time** and **increase capacity** in the system for those who most need to see a dentist:

> *“…of course, it will free up the dentist’s time to be able to see those people who he/she does need to see.”* P73 Phase 2

> *“I guess, for the patient, if the wider team can perform and undertake various procedures or interventions, then you might be able to see one of them quicker than perhaps another one.”* P71 Phase 2

##### 4.2.2.3 Potential challenges of skill-mixing and a whole-team approach

In addition to the potential benefits, participants also reflected on some **potential challenges**, or downsides, of this approach. Some felt that it could lead to a **loss in continuity:**

> *“Loss of continuity…I think if maybe you are seen by different people, so there’s that tendency or possibility that maybe you may not have a continued care.”* P21

Others were concerned that there might be a **lack of communication** between the different professionals, **meaning that things get ‘missed’ or ‘lost’:**

> *“If you split things out, you’ve got to ensure that there is that communication between all the team and that that communication is timely…you do end up with things [going] missing, things are missed because there’s ‘too many chefs then’, you have to be very careful you are clear on who’s doing what, and what communication matters between them, if something..”* P73 Phase 2

Some participants felt that this approach might lead to more **fragmented care** and **more appointments;** and so, taking up more time:

> *“Would it mean you’d have more multiple visits because only one person can do one thing and the other person does the other thing?”* P76 Phase 2

> *“…it can sometimes result in having to book a couple of appointments rather than getting the treatments done in one visit.”* P2 Phase 2

#### 4.2.3 Urgent and emergency dental services

Phase 1 and Phase 2 participants reflected on *what* they would do in urgent or emergency dental situations (including where they would go and who they would contact), *how* they would decide whether to access urgent or emergency dental care (EDC), preferences for services, and information needs to improve understanding.

##### 4.2.3.1 Uncertainty on how to access Emergency Dental Care

Many participants reported **uncertainty on who to contact or where to go if they had an urgent or emergency dental situation:**

> *“Oh, now you’re going into a very complicated area…I mean, that’s a whole can of worms…emergency dental services is still not clearly defined or understood by the patients and members of the public, and I suspect even the dental professions themselves between one dental profession and another dental profession.”* P75 Phase 2

> *“That is part of what I was describing, there are no more information as to how people would go about that. Because yeah, I do not know how to access it.”* P12 Phase 1

Several participants reported that they would **contact their dentist in the first instance:**

> *“Contact your, your dentist that you registered with.”* P76 Phase 2

> *“Well, I feel I would just go through the normal booking processes.”* P30 Phase 1

For some, this was due to the **trust and relationship** that they have with this known dentist:

> *“my first port of call is always going to be to my dentist because I have much more trust in my dentist. And then I will get referred from my dentist to someone else.”* P75 Phase 2

Others viewed this process as a **signposting opportunity t**o the correct EDC services:

> *“So, the reason why I say we need to contact our dentist is because our option may not be perfect. So, I think the dentist is the person that is in a perfect place to give you legit guidance on what you’re supposed to do at that time.”* P22 Phase 1

Only a few participants said that they would **use the NHS 11 service** to access EDC, but this was usually if the dental practice was closed and used as a back-up option:

> *“…if it is out of hours, if it is on the weekend, or when my dental practice is closed, then I will go into 111 services to get access to emergency dental care.”* P75 Phase 2

Some participants said that, depending on the severity, they would **call 999 or attend A&E:**

> *“Well, for me, I think our first point of call would be A&E at the hospital, and from there, they would… I think from there they would direct us to the actual section that we are supposed to be at.”* P52 Phase 1

##### 4.2.3.2 Deciding if something is a dental emergency

Participants reflected on a **range of factors that would help them to identify if their situation was a dental emergency**. These included, constant bleeding, severe pain that cannot be managed by regular pain killers, post oral surgery complications, unexpected injury (e.g. life threatening accidents, domestic violence), where there is infection and a risk of sepsis, and if they feel that they needed support or treatment within less than 24 hours.

However, some participants reported feeling **unsure about what an emergency is:**

> *“And also I don’t really know what, what constitutes an emergency.”* P74 Phase 2

Some participants report previous experiences of accessing EDC, which helped them to know where they should go and when (see Table 2).

##### 4.2.3.3 Better education of EDC and what constitutes a dental emergency and where to go

Participants reflected on their educational needs so that they could better understand what constitutes a dental emergency and where they should go to access this:

> *“We need a bit of a campaign to say this is what you do with a dental emergency. But also, what is an emergency? So, if I’ve got toothache, that can probably wait until tomorrow. Or I might take some paracetamol and it might go away.”* P74 Phase 1

##### 4.2.3.4 Preferences for what emergency dental services should look like

Many participants reflected that **location and distance to the service** were important to them. Most participants said that they would be happy to travel between 10-15 miles, or drive approximately 30 minutes, to access emergency dental care.

> *“I’m expecting not to travel so long in the case of an emergency because it’s called emergency, anything can happen.”* P13 Phase 1

Some noted that lack of transportation and long distances would be a barrier to accessing the service:

> *“If you haven’t got transport, then obviously, that’s a massive, massive issue because it’s how do people get from A to B, even if they want A&E, if they have to get a taxi or something, then it’s the cost implications for the taxi.”* P8 Phase 1

**Greater awareness of services and clarity on how to access was also important to participants**. They stated that there should be more information available about *how* to access EDS so there is a ‘known route’ and the systems used to make the appointment should be *accessible* and help people to navigate it:

> *“…I also feel that people should be educated, like we should create awareness whereby people are learning about the emergency dental services. That is going to help people seek immediate care. And [know] how to handle dental emergencies.”* P35 Phase 1

> *“…lack of awareness, because a lot of individuals might not be aware of their existence, or anything that has to do with emergency, so them lacking knowledge can also make them to be ignorant about what is happening to themselves, or maybe people around them. So we should try to create more awareness and teach people on emergency dental services.”* P27 Phase 1

#### 4.2.4 Essential general dental services and mode of delivery

This theme emerged from Phase 1 interviews. Participants considered the general dental services they perceived as essential, different types of delivery model (e.g. registered dental practice or health hub), and frequency of access.

##### 4.2.4.1 Essential versus non-essential services

For most participants, routine check-ups (and associated treatments) and treatment of urgent issues (e.g. broken tooth, displaced filling) were essential services that should be provided by NHS general dental services. For some, regular check-ups played an important role in prevention of oral health issues:

> “Regular check-ups are essential, because we’re trying to move into a space with health, aren’t we, where prevention comes first. If people can’t get check-ups, then we could easily be creating problems down the line.” P6 Phase 1

Cosmetic, restorative and some cleaning procedures were not deemed essential services.

> *“unless there’s a medical need to have some cosmetic treatment, that should probably be a private provider rather than a health service provision.”* P24 Phase 1

##### 4.2.4.2 Delivery models

Participants discussed their preferences for different delivery models, including registering for a dental practice, health hubs and walk-in clinics. A range of preferences were reported across those interviewed.

Some participants were positive towards the idea of **dental care being provided via local health hubs**, stating that they offered greater **flexibility**, they would be more **efficient** (especially for people with multiple health conditions), they could promote **collaboration** between dental teams and other health professionals involved in patient’s broader health care, and offer a more **holistic service.**

> *“…speaking as a diabetic I have to go one place for eye screening, one place for foot screening, one place for my consultant, one place for my GP. You know there’s a lot of places to go to and if they could be collocated…it’s a bit easier and you’d get used to going to one place, and you know where the car parking is and all the bus routes.”* P6 Phase 1

> *“…there’s a place of collaboration…I think that’s, that’s what dental services in Wales should look like, because collaboration between dental professionals and other healthcare professionals, you know, to address some holistic health needs. So’ I think there should be a place of collaboration.”* P27 Phase 1

One participant noted that this joined up and co-located approach to dental provision with other health care services could help to **reinforce to the public that oral health is part of health**, as it is often forgotten about:

> *“So I think, [if] we’re building these big health parks and facilities across Wales, it’d be quite nice if dentistry could be pulled in…I think it’ll help people to see it as a health thing, because it can be forgotten about, can’t it? People go to their GP, but do they always go to their dentist?”* P6 Phase 1

However, other participants **preferred a registered dental practice model**. They felt this offers **continuity in care**, opportunity to build a **relationship with the dentist** which would allow them to have a better understand of the patient, and they were more reassured that the dental team would have **up-to-date and relevant information** about their oral health history.

> *“I think I would be more comfortable going to a practice that I know, I’ve been using it for a while and I know they have all my information, and you can just pick up where I left from my last treatment.”* P52 Phase 1

> *“Okay, I prefer going to a specific dental clinic where I’m registered because I believe they will know more, they understand more what’s my problem is and how to go about.”* P49 Phase 1

One participant noted that the continuity of care offered at a registered dental practice helps their autism:

> *“So for me, being autistic, it’s really important to have the same person and I value that a lot…I really value having my same dentist who knows me well, who understands me, understands my challenges.”* P9 Phase 1

**Walk-in approaches** to dental service delivery were also discussed by some participants, but usually as an **emergency option** and not as an option for general dental service provision:

> *“The thing is with a walk-in clinic that makes me think of the traditional model of A&E. So you walk in and you wait for hours. You see someone who you haven’t seen before, and you end up repeating everything. So, I think there’s a place for them in terms of the more urgent cases perhaps, but I prefer a model where I make an appointment, I go to see someone in a practice that I’ve either seen before or I’ve seen the partner in the room next door before.”* P6 Phase 1

> *“If it’s, if it’s an emergency, then maybe walk in clinics, yes. But I don’t think it should be like walk-in access for any sort of [dental issue].’* P43 Phase 1

##### 4.2.4.3 Recall intervals for routine check-ups

Participants described their preferences for **how frequently they access their dentist** and the ideal recall interval between check-ups. Many participants described that the **recall interval should depend on individual circumstances:**

> *“When it is necessary…when I’m saying ‘annual’ that’s, that’s taking an assumption that your teeth generally are okay. But if you know if you’ve got some ongoing problems, then, it might be quarterly or six monthly, depending on what the state of your teeth is in the first place.”* P8 Phase 1

> *“no one actually prays to visit the [dentist] every day. I just want a process whereby anytime I’m affected in any way in terms of my teeth, then I would…just want that access to be there.”* P13 Phase 1

> *“I mean I really, because when I was growing up it was drummed into us every six months. Now basically I suppose what I’m saying is whatever the guidelines are I’ll follow.”* P6 Phase 1

> *“So I guess, I mean, if it’s, if the dentist thinks you only need to be seen once a year, then that’s fine. It should be professional judgement.”* P3 Phase 1

Some participants noted that it was **more important for them to know that they could access the dentist when they needed it, rather than a set time frequency.**

> *“I feel like what would be important would be the knowledge that I can access it fairly promptly if I needed to.”* P24 Phase 1

Where participants did express a specific time-period for the recall interval, most participants said between **6 and 12 months.**

> *“So I think, I would probably think in terms of kind of the routine appointments, I think six monthly to a year seems fine.”* P4 Phase 1

#### 4.2.5 Digital technology and ‘teledentistry’

This theme emerged from Phase 1 and 2 data. Participants discussed their views on digital technologies and ‘teledentistry’ – or remote dental care delivery and consultation given to patients at a distance, utilising electronic communication and telecommunication technologies.

##### 4.2.5.1 Booking appointments online

Many participants reported that they would **prefer to book their dental appointments online** or using an application (on a phone or tablet). Participants were positive about the use of technology to support the booking process, stating that it offers convenience (allowing patients to arrange appointments and select appointment times that worked around their other commitments), reduces delays in getting the appointment (by selecting something that works for them in the first instance), and reduces additional work for staff and patients (e.g. making telephone calls to rearrange a date that is not convenient).

> *“Well I think that would be good, because like when we do things with British Gas to come and do the washing machine…we’d go in, and then take a time and a day which suits us. So that would be brilliant.”* P9 Phase 1

> *“I’ll put it this way that we are in a very digital stage where people are more conversant with using online platforms to do their thing, people are now close to the phone, even older people know how to navigate your way around the internet, and all of that. So yeah, I think if I’m to book an appointment, then definitely it should be online.”* P12 Phase 1

Whilst the large majority of participants preferred online booking, most also recognised that **other options should still be available** (e.g. telephone) as some groups might be **disadvantaged by an exclusively online approach**, including elderly patients, rural patients with poor internet connectivity, and patients who are not digitally literate.

> *“And I think we need to be mindful that there are still a lot of people out there who don’t use technology. So for those, you know I’m thinking of my elderly parents, there should still be a facility to ring up and make a call…it can’t be all online yet, because we’re just not quite there.”* P6 Phase 1

> *“…booking it online and selecting a time and then getting a confirmation email would be the way to go, except that we live in remote rural Wales, with a terrible internet connection and a dodgy mobile signal. So there’s plenty of the population that might be more comfortable with a phone call.”* P24 Phase 1

> *“some people will probably go for telephone still because not everybody is digitally literate. So it’s harder for them [in terms] accessibility.”* P43 Phase 1

##### 4.2.5.2 ‘Teledentistry’ – virtual communication and appointments

Participants discussed the potential role of ‘teledentistry’[10]. First, they reflected on **synchronous approaches**, which are defined as an interactive connection (phone call, video call, etc.) where providers and patients share information in real-time. Many participants felt that video or telephone appointments could play a role in providing some initial advice and triage whether an issue required face-to-face follow-up, which in turn might save time and be more convenient.

> *“…there’s a lot of benefits in that…if you’ve got a broken tooth, or if you think you’ve got a filling that’s come out, you can stick a webcam to where you think the problem is, and the dentist can make a visual assessment there and then. It might be that they can then do a risk assessment as to whether you should be going in within the next 24 hours, 48, 36, or whatever.”* P8 Phase 1

> *“I can actually show the dentist. So we’re saving time, saving me to have to drive…I could show my mouth, [they] could see what’s going on, you could also see my physical appearance if I’m in pain.”* P9 Phase 1

Some reported that virtual appointments have **benefits for people living in rural communities**, some distance from the dental practice, particularly in emergency dental situations.

> *“As a positive aspect, it is very good if you don’t have access to dentists and dental hospitals. And if you are unable to, if you live far away from the clinics, then it’s a very positive thing, especially for a dental emergency.”* P1 Phase 1

Others discussed the role of **asynchronous approaches**, defined as remote, non-real-time communication between providers and patients, allowing each to access a platform when and where they want (e.g. sending emails or completing a form on a website). Participants felt that this would be an efficient way to communicate with the dental team and to check if a face-to-face appointment was required.

> *“…that could be very helpful, especially if there’s a long wait [on the phone] to get an appointment…know my GP practice, you can fill out an e-consult form and send images, more information and request a phone call from the GP. Something similar I think would be really useful.”* P5 Phase 1

> *“I think it, it’s positive. You fill out a few questions, including a question about what you would like the dentist to do, which almost gives you some ownership.”* P5 Phase 1

Despite the overall positive attitude towards ‘teledentistry’, most participants **acknowledged that its potential role was dependent on the situation**, and there were circumstances when it would not be appropriate:

> *“I think if the case is not severe, then it is fine. If it is severe, they will have to see you physically to know exactly what you’re talking about; to have a way to explain it better.”* P37 Phase 1

Some participants reflected on experiences of virtual appointments in other health care settings; most stated that these were positive experiences. However, whilst many participants felt teledentistry would be appropriate in some circumstances, **some participants did not feel that virtual appointments would work in the dental setting**.

> *“Not personally…I know what a virtual appointment is and what that entails and everything to do with it, I just don’t think it would work in dentistry.”* P7 Phase 1

> *“…it’s not like a GP where you can go and talk to them, [the dentist] needs to look at your teeth. I can’t see how that would work, if I’m honest, unless you had some very good software that could look in the mouth.”* P6 Phase 1

Participants had concerns that virtual appointments might miss more subtle signals that could only be picked up face-to-face, whilst others had concerns that the technology might not support this:

> *“You know number one, patients may find challenges with technology, such as the poor internet connectivity, or lack of access to suitable devices to support effective communication. So that has to be the number one issue with virtual appointments.”* P27 Phase 1

> *“I’m gonna give an answer based on what I experienced coming on this call, you know, connection might be an issue sometimes. And, and proper examination might be an issue virtually, and there might be defects in giving out diagnosis, a proper diagnosis.”* P28 Phase 1

#### 4.2.6 Self-management of oral health

This theme emerged from Phase 2 data. Participants reflected on the role that the public should play in looking after their own oral health, the importance of education to help support better self-management, ways to deliver this education and support to the public, and potential barriers to and concerns of being more engaged in self-management practices.

##### 4.2.6.1 Patient responsibility for oral health

Participants discussed the role that the public should play in their own oral health. Many participants felt that **individuals had a responsibility to look after themselves** and be engaged in looking after their oral health to prevent dental issues from occurring:

> *“Patients should be proactive and should be taking care of their own teeth. It’s not down to the dentist to tell them to brush their teeth twice a day for two minutes, is it? It’s quite a simple thing to do, keeping an eye on your own gums and your own teeth to see if you’ve got any problems and seeking help as soon as you have any problems.”* P73 Phase 2

> *“I think yes, perhaps we can be given information by some of the health care professionals…but I think, for a lot of people, there is an element of autonomy and self-awareness that does need to be brought into this.”* P73 Phase 2

When discussing this, some participants compared it to the ‘normal’ level of patient responsibility in other areas of health; one person noted that **oral health is sometimes seen as an ‘after thought’:**

> *“We need to let the public know that they have a responsibility to look after themselves as well…I think everybody has a responsibility to look after themselves, whatever their circumstances are. It is very important…teeth seem to be an afterthought…”* P72 Phase 2

> *“I think it’s the patient’s responsibility primarily to look after their own dental health…obviously we all brush our teeth and things like that, but it’s important that you look at your diet, which is the same for your general health as well. So why treat your teeth any differently? If you want to have a healthy diet for your general health, then you should also really consider your diet regarding your dental health.”* P174 Phase 2

##### 4.2.6.2 The importance of raising awareness of prevention and providing education

Whilst many participants felt that the **public should play an important role in prevention of oral health issues** and have **greater responsibility** for this, most also felt that this cannot be achieved without **greater awareness** of the importance of prevention. Some felt that current awareness and knowledge of oral health and dental services is limited:

> *“I think it is a bit like an iceberg. Patients only understand 10% of what dental services actually mean…90% of the dental services is not as understood by patients, even simple things like self-management, you know, the fact that there is a way of brushing your teeth, there is a way of looking after your oral hygiene, there is a way of managing your oral health.”* P74 Phase

Most of the participants who discussed this topic felt that there is a **lack of, or absence, of appropriate education for the public**. If this was addressed, it would help people to become more engaged in their own self-management:

> *“So, it absolutely is the responsibility of the patient and the parents. But…we need a lot of education before the responsibility kind of gets switched on again for some people… we’ve got a bit of a gap to build a bridge there I think, to try and get the education out.”* P74 Phase 2

> *“I think it’s about the information actually. I think it’s just literally people being aware of it. I guess everything from the sugary foods that we eat and the impact of that on oral health or the things that we drink, perhaps not taking two sugars in a coffee and only taking one or none ideally. Then I guess there’s the information about teeth cleaning and how to do it and making sure that we remember gum health as well, which is probably quite overlooked.”* P73 Phase 2

One participant felt that **raising awareness** of the importance of prevention would be a gateway to **increased engagement in active self-management behaviours:**

> *“once people have got awareness, they’ve got a little bit of understanding of how to do the self-management, they will then engage with the dentist. Now I hear, ‘I can do this and this. Can you please teach me how to do my brushing? Can you please tell me which fluoride toothpaste shall I use? Or how shall I do my…? How do I take care of my teeth?’ So they will engage with the professional in a meaningful way.”* P75 Phase 2

Another participant felt that **better education could help people be less reactive to oral health** issues, and more involved in preventing issues from happening:

> *“…they don’t think of the teeth as being a problem…until something goes wrong with your teeth, you don’t seem to worry about it, do you? Maybe an earlier sort of education system whereby you explain the need for people to look after their teeth because it can lead to so many other problems.”* P72 Phase 2

##### 4.2.6.3 Providing education to raise awareness of and engagement in self-management – best ways to achieve this

Participants described who they felt had the **responsibility of providing education** and **ways in which this could be provided.**

###### Welsh Government / NHS Wales Campaign

Some felt that the Welsh Government and NHS Wales had an important role to play in developing these resources and distributing them. They proposed that a **national level campaign** would be an effective way to **raise awareness** of the importance of prevention and being involved in self-management of oral health:

> *“…obviously then there’s the broader picture if you like, where you utilise the Welsh [Government’s] communications processes. Local health boards, the way they communicate to the public in their own local areas about other issues. it is important to communicate [to the public] whatever it is you want them to do.”* P72 Phase 2

> *“in terms of self-management, first of all, you need to make people aware that there is a need for, so examples of COVID examples…examples of smoking…the public media and public awareness was first done. So that’s the awareness part. Until you create that awareness, no one else is going to take an interest in it, [then] it is in front of people’s head to be aware about self-management…”* P75 Phase 3

###### Dental Team

Whilst a national campaign would be useful for raising awareness of the importance of self-management, some participants felt that the **dental team played a critical role in providing more education**:

> *“There needs to be better engagement between the dental services and the members of the public. I found out recently…that there are still things that I could do…I am not brushing effectively. So even though I consider myself informed, fairly educated, fairly intelligent, I only found that six months ago that I haven’t been brushing my teeth properly.”* P75 Phase 2

> *“The dentist could be playing a bigger role as well in these discussions around preventative measures, as that is the first team member that most people, when you go to the dentist for a check-up, do come into contact with. So perhaps they have a role to play as well.”* P134 Phase 2

Some participants reflected on **previous experiences**, where they felt that they were engaging in their own oral health and seeking information from dental teams, and on the **potential benefit of receiving increased education from the dental team:**

> *“ thinking of my visits over the years, I’ve actually been the one to ask the dentist about recommendations, to recommend products that might be good for my oral health. They’ve not expressed to me or discussed with me, trying to inform me about how to take care of my oral health, but I did discover that if you do bring it up, I think most times they are more than willing to engage around this discussion.”* P104 Phase 2

> *“I don’t think that’s something that a f dentist has ever asked me throughout my life. But I think it would help get that buy in from myself…if you set a goal then you want to achieve that goal and so you might be more inclined to make sure you routinely keep up the good practices. Because at the moment, you go to the dentist, you get the check-up, you hope everything is okay and then, ‘see you again in a year or two years’.”* P174 Phase 2

###### Schools and a school dental service

Several participants felt that schools would be an ideal route for the education, providing the skills and knowledge needed, and helping to **instil the importance of prevention at an early age:**

> *“I’m a former primary school teacher. And I very much believe that children, and young people, should be skilled up and have the knowledge and understanding of some of these health things, including oral health - so perhaps build that into the school curriculum or have lessons about oral health, dental health, and really start getting into prevention at a very early age.”* P73 Phase 2

Some felt that schools were the ideal setting but were also conscious of the additional workload on teachers. As such, they felt a **‘school dental service’** would be ideal. Not only would this be an opportunity to check children who do not currently present to dental services, but also a way to provide education at the same time:

> *“in these times, that would be a very good service to reinstate, with families not being able to access NHS dentists very easily now for various reasons. At least these children would be checked, and any problems then referred to a dentist, through a school system…their role could be to educate the children as well. You can’t expect teachers with everything they’ve got going on [to do this] … I think there would be a really important role now for a school dentist to visit each school occasionally and as well as checking the children’s teeth, educating them as well.”* P173 Phase 2

Others felt that the school-based approach would not only provide education to children, but could lead to a ‘knock-on-effect’ where **children share this information parents and others at home:**

> *“I think young people can handle it, and will probably go home and tell whoever’s at home, you know, we had a lesson about teeth and dentists and how to clean our teeth today. And then hopefully there will be that knock on effect, not only for the child, but maybe for whoever’s at home as well. So you could be killing two birds with one stone in terms of preventative activity for the teeth.”* P73 Phase 2

###### Community outreach

Some participants suggested community outreach programmes, for both the **broader population** and specifically for **harder to reach groups:**

> *“In some local communities or in some places, it’s very hard for them to access this care. And I know there are people, there are some people who are not even aware of this care. So I think my suggestion is that awareness should be created for people who are maybe in the rural areas, to know about this care.”* P72 Phase 1

###### Inclusive options

Some participants reflected that regardless of the route to provide the education, the needs of different people should be considered so that they receive the **information in an inclusive way that works for them:**

> *“You educate people in the best way for them as individuals, you know, because we’re not all the same. Some people can’t read, [some] are blind and deaf, how do you do it then. You have to look at it in a holistic way.”* P72 Phase 2

##### 4.2.6.4 Barriers to engaging in self-management and prevention

Participants discussed potential barriers to the public becoming more engaged in the oral health self-management and taking a larger role in preventative care. Several participants described **costs as a key barrier:**

> *“Look at the cost of living crisis right now, there are families who have to decide whether they’re gonna buy toothpaste or whether they’re gonna buy food. And then when you look at things like flossing, mouthwash…lots of people can afford that but there’s also lots and lots of families who cannot afford those products. So, it’s not just the individual’s responsibility in terms of this.”* P174 Phase 2

> *“…they may not have the money…there’s no point telling someone, ‘you need to brush your teeth five times a day’, when they can’t even afford to buy a brush or toothpaste.”* P75 Phase 2

Participants discussed the importance of education at length (see previous sections), but they also saw **lack of awareness and education** of best practices as a **key barrier to engaging in effective self-management:**

> *“There’s no point telling someone to brush five times a day, if they don’t know how to brush, and they don’t know how to do to clean their teeth.”* P75 Phase 2

> *“So one of my bugbears, there’s no communication in certain circumstances, [they] don’t actually let you know what it is they want you to do.”* P72 Phase 2

For some, **cognitive capacity** might be a barrier to engaging in self-management of their oral health:

> *“I appreciate that for some people that may not be possible, like people with learning disabilities or a mental health problem or Alzheimer’s…there a certain conditions or areas when people might not be able to have that level of autonomy, and to look after their own oral or dental health.”* P73 Phase 2

**Early experiences** were also considered by some participants as a barrier to preventative oral health behaviours:

> *“I think education, most of it is our experience when we’re young. A lot of people don’t live in households where they have that kind of supportive [role], parents who are encouraging them or nagging them to do their teeth, so they don’t get into that habit. They don’t understand why it’s important and that can lead to kind of problems later on in life.”* P174 Phase 2

Some participants note that **oral health can take less of a priority when patients are experiencing other health issues:**

> *“…that might be the bottom of their list…thinking that logic through, especially if they’ve perhaps got more severe medical health problems, cancer for example, could be anything…having a bit of a toothache, that might literally drop down the bottom of the list if you’re receiving palliative care…”* P73 Phase 2

Several participants recognised that the **barriers to engaging in self-management were complex** and exist at different levels, and does not only include individual factors:

> *“So this isn’t a simple solution in terms ‘you just need to brush your teeth twice a day, there’s a whole lot of complex factors that contribute to why people do the behaviours that they do. So just focusing on individual behaviours isn’t gonna be enough to shift to a preventative society.”* P174 Phase 2

##### 4.2.6.5 Concerns over increased self-management

Whilst most participants believed patients should be more involved in looking after their own oral health to prevent problems and were positive about the idea of greater self-management, some participants raised potential concerns. This included the worry that improved self-management would result in fewer people presenting to dentists, which could result in **important issues being missed:**

> *“…they do everything they’re told to do, they think they don’t need to go and see a dentist. And actually, they do need to see a dentist because there’s something that’s brewing, in the pipeline that they aren’t aware of, that the dentist can pick up on much earlier to stop them losing their tooth or whatever.”* P75 Phase 2

There was also concern that if more people take care of their own oral health, and fewer people present to dentists, it could **impact supply and demand, resulting in NHS dental services being cut:**

> *“…dentistry is almost a supply and demand profession to some extent. So if people are taking more care of their own oral health and there’s a lot more prevention, at the end of the day, maybe not many people might need to go to the dentist, or might not need to go for such severe problems, which would then mean a lack of need for dentists…so it could be a workforce implication there.”* P73 Phase 2

Other concerns focused on **increasing inequity in oral health outcomes**, with some people being asked to engage in self-management who might not have the necessary motivation, knowledge, and skills to do so:

> *“Another downside could be that some people may not want to or be able to take care of their own oral health. And that then starts to make the gap between those who can and those who can’t, those who do and those who don’t, even wider - that kind of inequity.”* P73 Phase 2

#### 4.2.7 What matters most to the public – key priorities for dental services in Wales

Our final theme emerged from Phase 1 and Phase 2 data. Participants reflected on **what mattered most to them** – **or their key priorities for dental services in Wales.**

**Six key priority areas were described** (see Figure 6).

**Figure 6:**
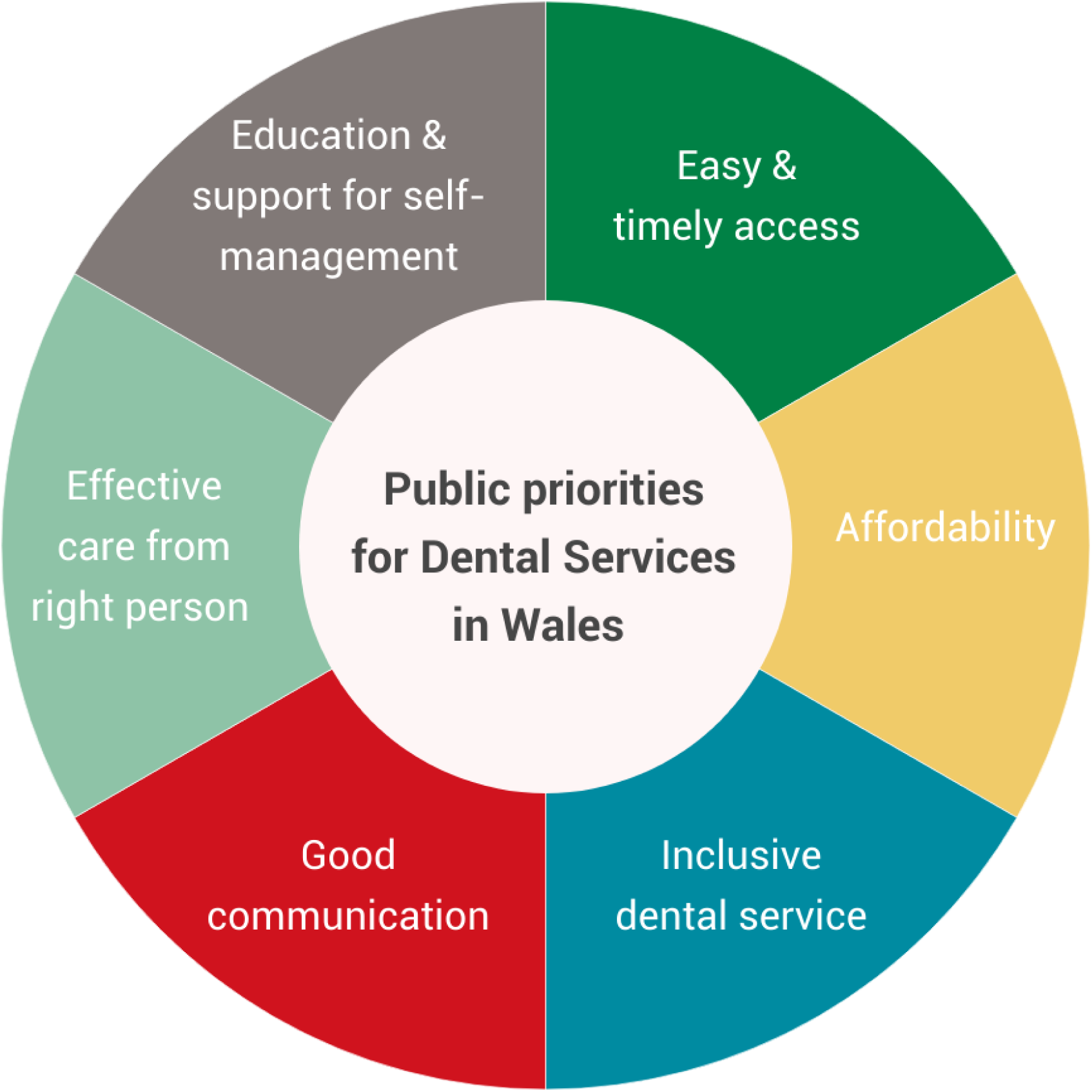
Six key priorities for dental services in Wales

##### 4.2.7.1 Easy and timely access

**Easy and timely access to dental services** was the top priority stated by most participants.

> *“Well, I think the most important thing is to have a service which is available…that provides the right level of care in a professional manner, which is easily accessible. Those three things will be my main criteria.”* P72 Phase 2

> *“The thing that matters most to me is having access to a dental practice.”* P73 Phase 2

Many participants report that **being able to register with an NHS dentist when you are eligible to** is the thing that matters most to them. However, they describe experiences contrary to this need:

> *“You could be on a huge waiting list just to get an NHS dentist, never mind being seen. You can’t even get on their list. So I think there should be sufficient NHS dentists, or sufficient services provided to people. You shouldn’t have to pay a private dentist to be seen [when you’re eligible for NHS services]…it’s very difficult I think.”* P6 Phase 1

> *“Because we, we moved into [Placename] two years ago, and couldn’t get on a dental list anywhere, not even within a 20-mile radius probably.”* P24 Phase 1

Others reported concerns that access to NHS dentists is and will continue to become more challenging, as more NHS dentists move to private practice:

> *“Well, I am quite concerned, if my dentist decides she wants to give up the NHS contract, I don’t know quite what I’d do then…it’s not insignificant what you have to pay.”* P33 Phase 1

In addition to being able to access and register with an NHS dentist, **timely access to the service when you need it** was also important to participants:

> *“The crucial bit when I say accessible it’s to be able to get an appointment on a fairly timely basis…if you have toothache, to be able to get an appointment, ideally, the same week, or couple of weeks. Obviously, we’re a long way from that ideal at the moment.”* P24 Phase 1

> *“I think being able to get an appointment, you know, when you need one…that’s something I’ve found problematic before in the past, whether it’s been NHS or private, being able to get an appointment when you need one.”* P45 Phase 1

> *“And what’s more important to me is how timely the care is being delivered.”* P53 Phase 1

When discussing ease of access, participants also referred to the **ease of scheduling the appointment**, or the appointment booking processes, as something that was important to them:

> *“I think it should be easy to access and not so stressful. In the sense that a person who wants to use or get the services, and then you are not to be stressed.”* P38 Phase 1

> *“…the kind of services I would expect to receive would start from scheduling an appointment, there should be a well-organised system of scheduling these appointments.”* P28 Phase 1

##### 4.2.7.2 Affordability

Many participants stated that **affordability of dental care was an important priority.** Lack of affordability was discussed as a **significant barrier** to some people seeking dental care:

> *“If you look at their individual circumstances, can they afford to pay? There shouldn’t be a barrier in some respects in coming to dentists because they can’t afford to pay.”* P72 Phase 2

> *“Some of the costs are, in my opinion, a bit extortionate. Obviously, but then I’m not a dentist, I’m not running a practice. I don’t know what his overheads are or anything else. So I think it’s extremely difficult if someone’s on benefits, it’s difficult to pay for something like dental services. You get the free NHS check, but then all the add-ons, like the false teeth, the other bits and pieces, the polish, you know, scale and polish, it all costs money. Sometimes people can’t afford that money”* P74 Phase 2

In relation to costs, participants also noted that there is a **lack of clarity about why there are charges for NHS dental services**, but not for other NHS services:

> *“the affordability is a massive thing as well. But I guess I don’t really understand why you, you have to pay for dental care, whereas, you know, most other care on the NHS is, is free. So a little bit of understanding why that is the case is needed”* P174 Phase 2

##### 4.2.7.3 Inclusive dental services

Participants emphasised the importance of an **inclusive NHS general dental service** for all who are eligible to use it; on that is fair and non-judgemental. Some participants noted that the most important thing to them was an inclusive service that **treated people from different ethnic backgrounds fairly and equally:**

> *“what’s more important to me is to treat me like other patients who are…you know sometimes, we black [people], we experience a lot of forms of discrimination sometimes…that is my expectation, my expectation is to be treated the same way every other person is being treated.”* P49 Phase 1

Other participants discussed inclusivity in relation to **financial challenges** faced by some people:

> *“I think our dental services should be accessible for everyone in Wales, most especially, people having financial challenges to be given an equal opportunity to treatment in Wales.”* P44 Phase 1

Some participants discussed how the service could be more inclusive for people who are neurodivergent or have additional learning needs:

> *“I am autistic and sometimes it’s having a dentist who like understands that some people might struggle a little bit more than others. So having someone compassionate is really important as well.”* P4 Phase 1

> *“You see posts online going ‘does anyone know of any dentists that are sympathetic and good with like autistic children or children with additional needs’…[we need] a way to allay people’s fears, and not feel like they’re being judged, whether as a parent, an individual, or whatever it is.”* P45 Phase 1

Inclusive access for people who are **employed full-time** and are unable to access the dentist during the working week and for **people living in rural areas** was also important:

> *“There should be accessibility to all demographics…regardless of social economic factors and geographic location, especially for those living in the remote areas, there should be access to dental services. That’s it.”* P27 Phase 1

##### 4.2.7.4 Good communication

**Good communication** between the dental team and patients was a key priority for participants, and was seen as the **foundation of good dental care:**

> *“It’s all about the relationship and the communication environment that you’ve created with your dentist.”* P33 Phase 1

> *“I mentioned earlier on about proper communication with each patient, and I’m always talking about the patient because delivery should be very patient centred.”* P28 Phase 1

Participants reflected on previous instances of poor communication, which resulted in negative experiences, to reinforce their views on its importance:

> *“Just being kept in the loop about basic things…they started charging upfront for things in advance, I wasn’t notified about that…that’s quite an important thing. So why did that happen? Was it just something they put up in, in the practice that you’d only be aware if you actually saw it, there was nothing on their social media. It’s channels of communication, isn’t it? I think sometimes that can be the biggest issue, but the most simplest one, just looking at ways that you exchange information.”* P45 Phase 1

##### 4.2.7.5 Effective care from the right person

Another priority for participants was having confidence that **the person you are seeing was qualified** to carry out the tasks **safely and effectively:**

> *“And then I want the best person I can possibly get with the best training and the best support as well, within that practice.”* P72 Phase 2

> *“For me, that’s important…it’s the sort of safety aspect of it - that the people that are doing the work, the treatment, that they are qualified, they have got all the relevant registrations and rest of it. So yeah, I’d probably say price, time and safety would be the three most important things.”* P71 Phase 2

Participants also noted that it was important that they could **access the right person for the right task:**

> *“Being able to access the right person, you know, for the treatment that you want.”* P45 Phase 1

> *“I’d be happy enough to meet with whoever could give me the information that I wanted at the time… all I’m interested in is getting the right results.”* P8 Phase 1

> *“I don’t mind who I see as long as they’re qualified for what needs doing, you know?”* P3 Phase 1

##### 4.2.7.6 Better education and support for self-management

The sixth key priority discussed by participants was better **provision of education** and **support from dental teams** to enable them to take a more **proactive role in oral health self-management.** This was also discussed at length as a separate theme (4.2.6 Better support for self-management), but participants explicitly stated it was also a priority for dental services in Wales.

> *“In summary, there should be more accessible [education] materials, healthcare services and dental personnel.”* P18 Phase 1

> *“Education on proper oral hygiene and the role and importance of a healthy be comprehensive, you know, accessible and preventive…it should be a place of education”* P27 Phase 1

Participants described a variety of routes for evidence-based education and support including via the dental practice (e.g. flyers to raise awareness), dental team (e.g. leaflets handed out during appointments), online information hosted by NHS Wales / Welsh Government (e.g. website where information could be accessed and dental care plans downloaded), social media (e.g. Facebook, TikTok, X to raise awareness and signpost materials), and targeted outreach programmes in schools, rural areas, and underserved communities.

## 5. Discussion

We explored what the Welsh public *understand* about NHS dental services, what they *think they could look like*, and their *key priorities*. Our in-depth qualitative study with 44 members of the public revealed that most participants: have *limited understanding of the dental team* and their roles; *lack clarity* on how and when to *access EDC*; are *supportive of skill-mixing* and multi-disciplinary approaches to their general dental care, but *awareness* and *education* are needed first; are *receptive* to solutions offered by *‘teledentistry’*, if this is delivered in an *optional* and *inclusive* way; and *would like greater education and support* to be more engaged in *oral health self-management*. **Six key priorities for dental services in Wales** were identified by participants:

**1.** Easy and timely access
**2.** Affordability
**3.** Inclusive dental service
**4.** Good communication
**5.** Effective care from the right person
**6.** Education and support for self-management

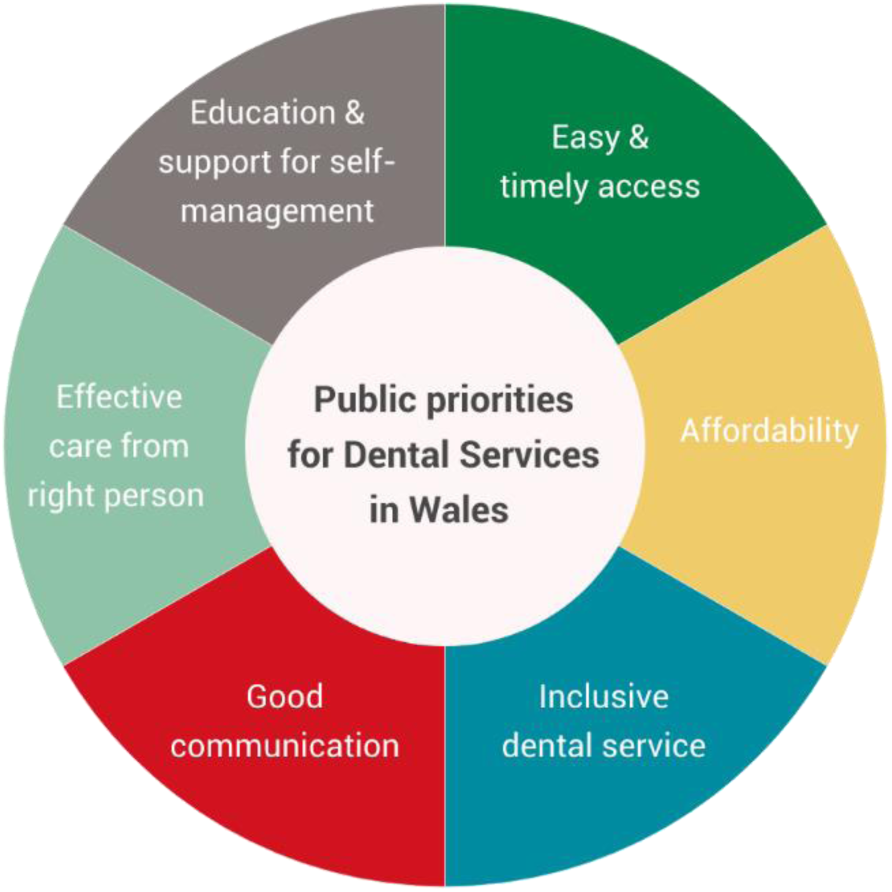

Participants were acutely aware and mindful of the widely documented pressures on NHS dentistry, including access issues[11]. Whilst they frequently used personal experiences to illuminate their points, including those related to access, we also found that the participants were largely solution-focused - reflecting on what their needs were but also making recommendations for how those might be feasibly met in NHS dentistry. This solution-based approach was reflected across all seven key themes that emerged (see Table 2) and the six key priorities outlined by participants (see Figure 6). Below we consider key discussion points linked to the key priorities for dental services.

### 5.1 Multiple and tailored delivery models – the role of risk-based recall, health hubs, and digital technology

When describing what they thought should be delivered as part of the GDS, **participants’ expectations were largely in line with current service delivery and provision**. Essential services included routine check-ups and treatment of urgent issues, whereas cosmetic treatments were not essential; importantly, no key gaps in services were identified.

Implementation of needs-led dental recall intervals is a key goal of the dental reform programme in Wales, [12] and the rest of the UK.[13] Despite being recommended in NICE Guidelines 20 years ago,[14] evidence suggests that many patients attend at 6-9 months intervals.[15] Other studies, including an independent evaluation of the reform programme, found that some patients have concerns about recall intervals over 12 months.[16, 17]

Length of recall interval for routine check-ups was discussed by our participants. Some participants said that the ideal interval would be somewhere between 6-12 months.

However, many of these also noted that there was no specific reason for this, other than ‘that is what they’ve always done’. Several participants, including some who reported a preference for a 6-12 month recall interval, stated that the interval should ultimately depend on individual circumstances and tailored to the patient. Reported benefits included freeing-up capacity in NHS services for those who need access, who either do not currently attend (e.g. people from underserved communities) or cannot attend (e.g. unable to register with an NHS dentist). These findings show **that participants were receptive to a risk-based recall interval that works best for them, decided together with the dental team.**

We also found that **participants were receptive to and expressed a preference for a range of delivery models.** Whilst some participants preferred the continuity offered by a registration model, others felt that the ‘health hub’ option offered many benefits: increased flexibility, efficient access when you need it, improved collaboration between the dental team and other health care professionals involved in a person’s care, thus providing a more holistic approach. Another that theme relating to ‘how’ dental services are delivered was the potential role of digital technology. ‘Telemedicine’ has been increasingly implemented as an alternative service delivery option in other areas of health care, accelerated in part by the COVID-19 pandemic, and showing effectiveness in making diagnoses, prescribing medications, providing timely care and increased convenience for patients.[18]

However, its adoption in dental care has been slow, due in part, to the practical nature of dentistry[19]. Despite this, **many participants were positive to the option of live virtual consultations** (e.g. video consultation with member of the dental team) **or digitally enabled interactions** (e.g. sending request forms and images via a website link), citing several benefits, including convenience, less time wastage in the system, and better access for those in rural communities. These views align with the findings of a validated survey that explored patient experience of virtual consultations in oral medicine during the pandemic; over 82% rated their experience as good or very good, and 69% preferred a virtual appointment for their next consultation.[20]

If this mode of delivery was used, the most important thing to participants was that a) it was *inclusive* and there was a *choice* to use the digital services, and b) that there were clear parameters regarding when they were appropriate, and *clear pathways to escalation* if a face-to-face appointment was required. There was also a **strong preference among participants for dental services to use and improve digital booking systems**; again, participants reported that these offer convenience (e.g. being able to make an appointment outside of working hours) and less time wastage (e.g. avoiding being sent an appointment that does not work, and then having to call the dentist to rearrange). As with all changes, this should be delivered in an inclusive way – offered as an option, but not the only route to making an appointment.

The alternative modes of delivery described by participants could offer a solution for three of their key priorities for dental services in Wales. Streamlined digital booking systems could support *easy access*. Risk-based recall, health hubs and digital consultations or interactions could improve *timely access*. Digital opportunities could also provide a more *inclusive* dental service to people living in rural communities and improve two-way *communication* between patients and dental teams.

### 5.2 Could improved dental team understanding underpin successful skill-mix approaches in dentistry?

The use of skill mix in NHS general dental practice has been a significant issue over recent years. Seeking to maximise the potential of the wider dental team, it is also presented as a potential remedy for workforce recruitment and retention and dental service access issues. NHS England made significant changes to the NHS dental contract in 2022, removing administrative barriers preventing dental care professionals operating within their full scope of practice.[21] Subsequent guidance[22] provides information on how to achieve this in practice, including dental therapists and dental hygienists providing direct access to NHS care where it is within their scope of practice. Increasing the use of skill-mix is also a key goal of dental reform in Wales;[15] allowing clinical teams to look after the health of patients according to need and freeing up dentist time for new patient access or those with more complex needs.

**Most participants in our study were very positive to the use of skill-mix.** Overwhelmingly, the view was that **it was less important ‘who’ they saw – what mattered most was seeing the right person for the issue.** They also perceived several *benefits* of the approach for patients, including improved patient choice and person-centred dental care that aligns with their preferences. Importantly, they also described dental team and system level benefits, including improved collaboration between the dental team, upskilling opportunities for other staff, improved dental team morale, motivation for people to join the workforce, and increased capacity in the system. Participants compared skill-mix in dentistry favourably with the approach that is taken in other health care settings. However, they also pointed out that if they were seeing someone other than the ‘lead clinician’ in other settings (e.g. general practitioner or consultation) they typically knew *who* they were seeing (e.g. physiotherapist, nurse, pharmacist) and *why* they were seeing them – they reported that they do not have the same understanding in dentistry.

**Overall, participants had a limited understanding of who the dental team members were and what their roles were.** Most participants were only aware of three members - dentist, dental nurse, and hygienist – often using the term ‘dental assistant’ to refer to all team members other than the dentist. Some said they knew that there are different team members who do different things, but they could not name them, and they do not know what they do, or if they can do similar tasks to the dentist. Despite being one of the key roles included in the NHS England skill mix guidance,[22] no participants were aware of the ‘dental therapist’ role. If we want the public to be *receptive to skill-mix* and booking an appointment with alternative team members to free up capacity and improve access, a critical first step is to *improve understanding of a) who the dental team members are and, b) what their roles involve*.

One of the six key priorities stated by participants was *effective care from the right person.* Participants discussed having confidence that the person you were seeing was qualified to carry out the tasks safely and effectively. *If the public do not know who is in the team*, the titles of the different team members, or the tasks that they are able to perform, *how can they have confidence that the person they saw was the right person?* If we improve public understanding of the dental team, which could encourage people to book/be receptive to being allocated appointments with other team members, this could help to address several of their key priorities, including *easy and timely access*, *good communication, and effective care from the right person.* This will also address a key goal of dental reform in Wales.

### 5.3 Awareness first, then education with support – improving self-management and knowledge of services

Our findings show that **most participants believed that patients have a responsibility to be involved in looking after the own oral health**. There was also a desire among many patients to be more involved in their own oral health self-management. However, **participants felt that current lack of awareness, education, and support were critical barriers to better oral health self-management.** Importantly, they believed that many patients are not even aware of the importance of self-management and taking a greater preventative role in oral health, nor were they aware of the things that people could do to achieve this. A recent qualitative study corroborates this, demonstrating a lack of awareness of prevention in oral health.[23]

Participants referred to building a ‘bridge for this gap’. As a starting point and an important baseline, there should be **increased efforts to improve awareness among the general population about the importance of prevention and self-management in oral health.** In line with our findings, Leggart et al (2023)[23] have also shown that oral health is often not a priority for patients and is lower down their hierarchy of needs. By raising awareness of its importance, our participants believe this might help patients to not see dental health as an ‘after thought’ to other health issues, and it might ‘switch on awareness of this responsibility’. Preferred routes for awareness raising were *national level campaigns* supported by Welsh Government and NHS Wales.

Once awareness raising efforts are in place, **good quality coordinated education must be provided to the public so that they know what they can do** to be more involved in their own self-management and prevent dental issues from happening. Quite simply, we cannot expect people to play a greater role in preventing oral health issues if they lack the information to do so. Importantly, this **should not only involve access to the information. It should be underpinned by patient-centred support**; helping the patient to understand the information whilst enabling them to use the prevention behaviours. Most of our participants do not recall having preventative or self-management discussions with a dentist.

A range of education and support routes were suggested by our participants. *Dental teams* would play a key role in providing access to these materials. However, *school-based* and *community outreach* programmes should complement this – these inclusive approaches were deemed particularly important for those individuals who do not (or cannot) currently present to the dentist, for various reasons. Some excellent examples of education resources already exist including a self-management guide for orofacial pain[24] and the preventative ‘Designed to Smile’ programme for families and young children aged 0-7 in Wales, focused in disadvantaged areas.[25] However, the key will be to enable *coordinated access* to these types of education, *making sure there are inclusive education options available for all NHS dental patients*, regardless of current attendance patterns, across the lifespan and addressing various needs.

It is important to note that whilst there was general positivity towards increasing oral health self-management, **some patients were concerned about the potential repercussions of patients taking a more active role in prevention**. This was underpinned by ongoing concerns about access and reducing dental services – ‘if we look after our own teeth, and access the dentist less, will we be in a Catch-22 situation where services are reduced further?’. Improved prevention and education to self-manage are key to delivering a person-centred dental service in Wales and was one of the six key priorities stated by our participants: ‘*education and support for self-management’*. However, any messaging around prevention and education must be carefully balanced to alleviate these concerns over supply and demand.

A final and related issue was the **lack of knowledge about what constitutes a dental emergency and where to access support.** If patients are expected to self-manage, they need more guidance on what constitutes and urgent or emergency dental problem, and what can be self-managed at home. Studies report that 380,000 patients attend UK general practice per year with dental problems[26] and patients reporting to A&E with dental problems accounts for 1% of attendees[27]. Whilst the reasons for attending are numerous and complex (including costs and availability), improved awareness and education could help to guide patients to the appropriate dental care pathways.

### 5.4 Strengths and limitations

This study has explored the views of a diverse group of participants, representing multiple health boards across Wales, ages groups, ethnic backgrounds, employment/incomes, and disability status. The qualitative methods have provided in-depth understanding into participants’ needs and their reasons for these, whilst allowing us to explore possible solutions to these. However, whilst we have recruited a relatively diverse participant group, we should acknowledge that there could be a potential bias in this self-selecting sample, mainly due to the digital routes used for data collection. Further, most participants report themselves to be digitally literate (see 4.1.5). Future work should also explore the views of those who do not typically self-select for research participation and also use offline methods for data collection.

## 6. Implications for practice and recommended priority areas

Findings of this study outline **patient’s key priorities for their dental services in Wales** (Figure 5) and could be used to inform dental reform.

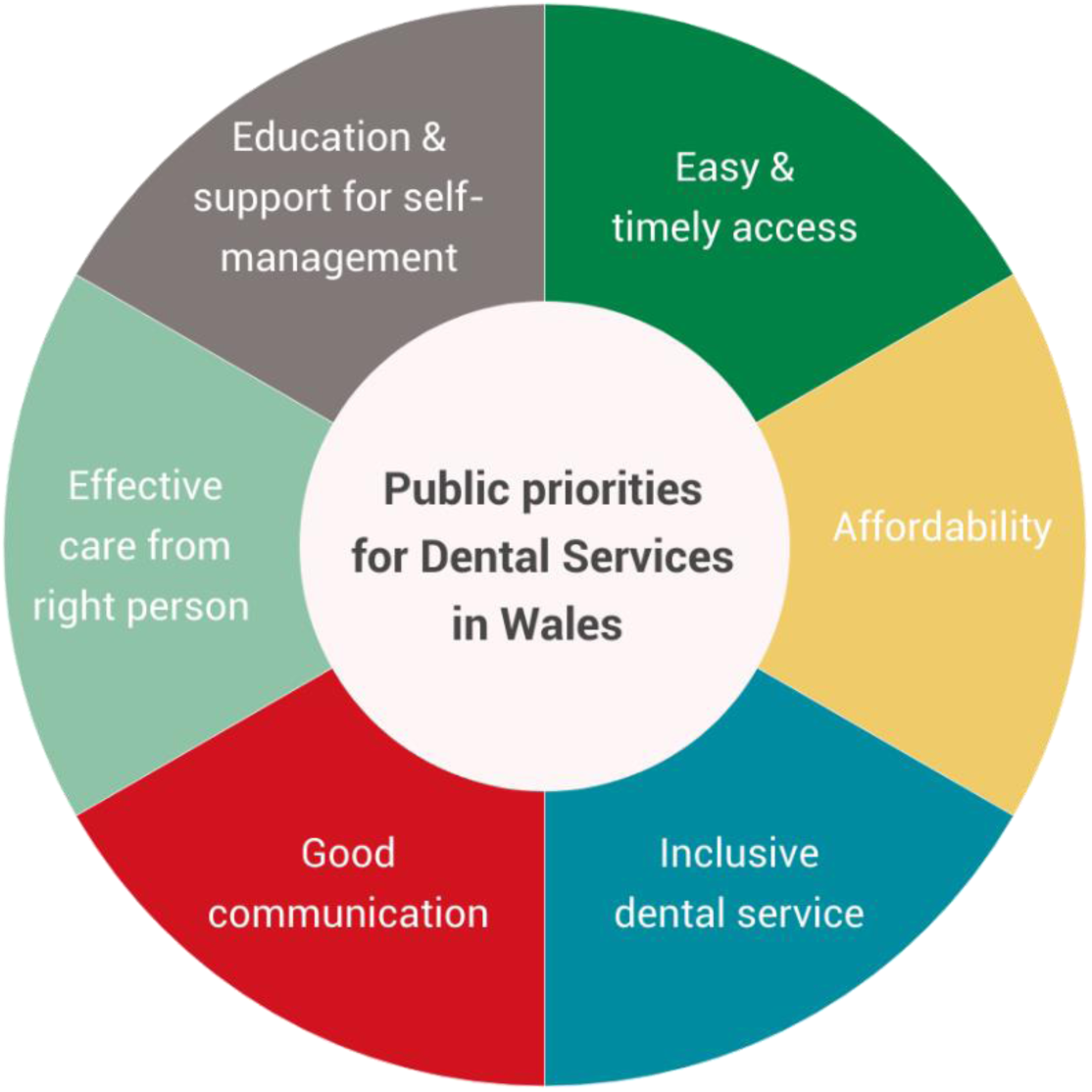

**Of specific relevance to the dental reform objectives are the following findings:**

- The public have *limited understanding of the dental team* and their roles,
- They *lack clarity* on *how* and *when to access EDC*
- The public are *supportive of skill-mixing* and multi-disciplinary approaches to their general dental care, *but awareness and education are needed*
- They are *receptive* to solutions offered by ‘*teledentistry’*, if this is delivered in an *optional and inclusive way*
- The public would like *more education and support* to be more engaged in oral health *self-management*.

Based on these findings, and the six key priorities, **we make the following recommendations for priority areas:**

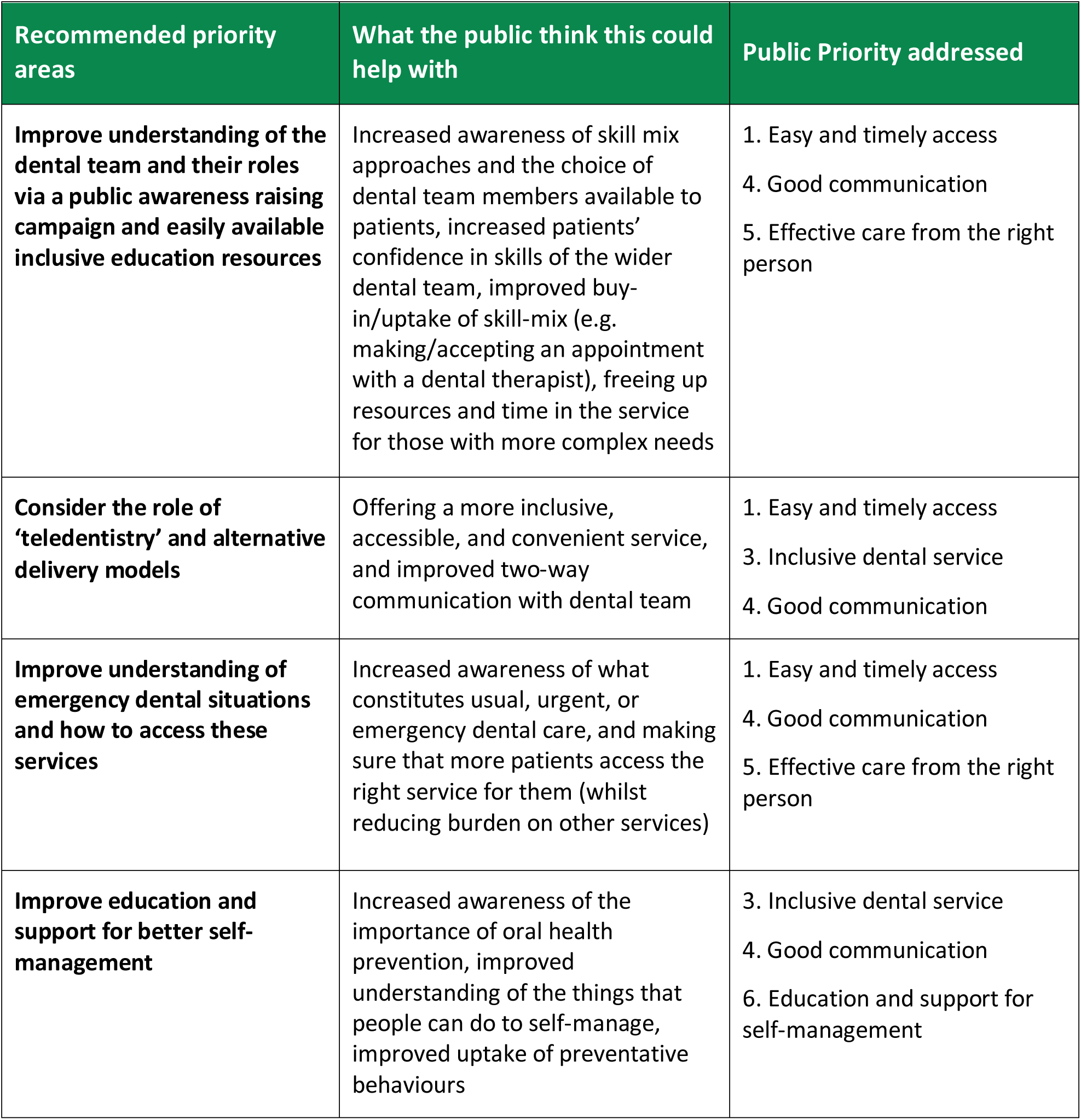

| Recommended priority areas | What the public think this could help with | Public Priority addressed |
| --- | --- | --- |
| **Improve understanding of the dental team and their roles via a public awareness raising campaign and easily available inclusive education resources** | Increased awareness of skill mix approaches and the choice of dental team members available to patients, increased patients' confidence in skills of the wider dental team, improved buy-in/uptake of skill-mix (e.g. making/accepting an appointment with a dental therapist), freeing up resources and time in the service for those with more complex needs | 1. Easy and timely access<br>4. Good communication<br>5. Effective care from the right person |
| **Consider the role of 'teledentistry' and alternative delivery models** | Offering a more inclusive, accessible, and convenient service, and improved two-way communication with dental team | 1. Easy and timely access<br>3. Inclusive dental service<br>4. Good communication |
| **Improve understanding of emergency dental situations and how to access these services** | Increased awareness of what constitutes usual, urgent, or emergency dental care, and making sure that more patients access the right service for them (whilst reducing burden on other services) | 1. Easy and timely access<br>4. Good communication<br>5. Effective care from the right person |
| **Improve education and support for better self-management** | Increased awareness of the importance of oral health prevention, improved understanding of the things that people can do to self-manage, improved uptake of preventative behaviours | 3. Inclusive dental service<br>4. Good communication<br>6. Education and support for self-management |

## Data Availability

All data produced in the present study are available upon reasonable request to the authors

## Acknowledgements

The authors would like to thank Dr Andrew Dickenson (Chief Dental Officer for Wales) and Dr Warren Tolley (Deputy Chief Dental Officer for Wales) for their involvement, time and expertise in the research process.

## Abbreviations

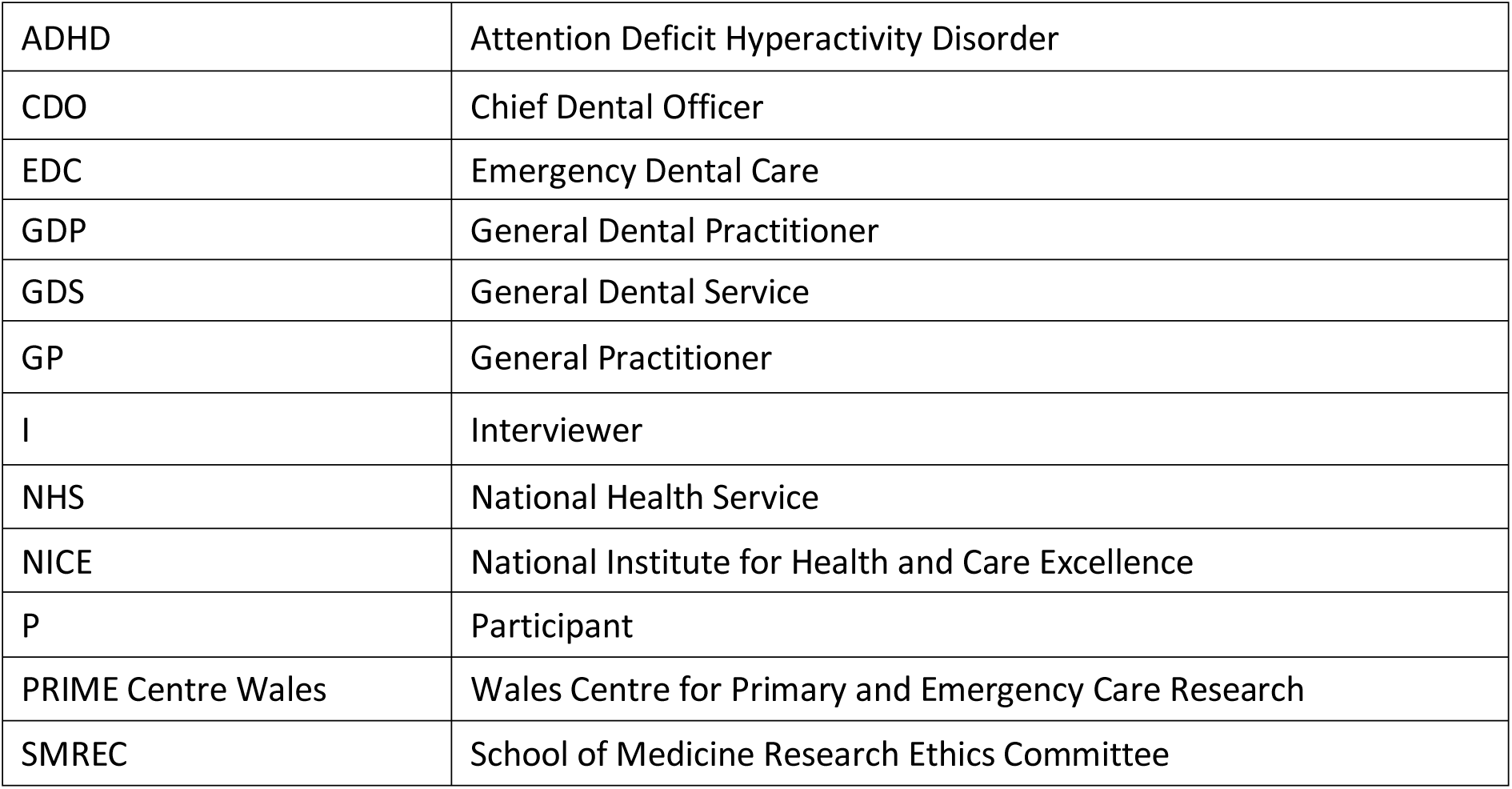

|  |  |
| --- | --- |
| ADHD | Attention Deficit Hyperactivity Disorder |
| CDO | Chief Dental Officer |
| EDC | Emergency Dental Care |
| GDP | General Dental Practitioner |
| GDS | General Dental Service |
| GP | General Practitioner |
| I | Interviewer |
| NHS | National Health Service |
| NICE | National Institute for Health and Care Excellence |
| P | Participant |
| PRIME Centre Wales | Wales Centre for Primary and Emergency Care Research |
| SMREC | School of Medicine Research Ethics Committee |

## Footnotes

1 In the exemplar quotes, ‘P’ denotes participant, and ‘I’ denotes interviewer.

